# Early Progressive Peripheral Airway Dysfunction after Allogeneic Haematopoietic Stem Cell Transplantation is associated with chronic Graft vs Host disease not BOS-0p

**DOI:** 10.1101/2023.11.03.23298073

**Authors:** Christopher Htun, Robin E Schoeffel, Sandra Rutting, Jacqueline Huvanandana, Cindy Thamrin, Alun Pope, Craig L Phillips, Matthew Greenwood, Victoria Pechey, Gregory G King, Paul D Robinson

## Abstract

**Background:** Current spirometric-based criteria for diagnosis of bronchiolitis obliterans syndrome (BOS) may miss early peripheral airway disease associate with disease onset. Multiple breath washout (MBW) and oscillometry offer improved sensitivity, but longitudinal changes occurring in allogeneic haematopoietic stem cell transplantation (HSCT) are unknown.

**Objective:** In this longitudinal study of HSCT survivors, we investigated changes in nitrogen-based MBW, oscillometry and conventional lung function, from baseline (pre-transplant), over 36-months, and associations with BOS Stage 0p, a spirometry-defined risk classification for potential later BOS development, and chronic graft-vs-host disease (cGVHD).

**Study Design:** Longitudinal observational study of allogeneic HSCT recipients from a single adult centre. All participants underwent spirometry, plethysmography, gas transfer capacity (DLCO), oscillometry and MBW at each study visit. Tests were performed pre-HSCT and 3 monthly thereafter over 36 months.

**Results:** 64 of 69 recipients recruited were included in the final analysis. Across the entire cohort, deterioration in acinar ventilation inhomogeneity (S_acin_) occurred as early as 90 days post-HSCT (0.224 z score change/month, p<0.001), prior to any significant change in spirometry or oscillometry. Progressive deteriorations in S_acin_ were associated with cGVHD status and grade but not BOS-0p status.

**Conclusions:** Early progressive peripheral airway dysfunction occurred following HSCT and was best detected by S_acin_ from MBW. Distal acinar ventilation inhomogeneity (S_acin_) deteriorated at an earlier stage than spirometry. Longitudinal deteriorations in S_acin_ were related to cGVHD, and independent of early changes in spirometry parameters. These findings suggest an important role of the lung in cGVHD and provide important evidence to support future studies examining the prognostic utility of MBW in long-term monitoring of HSCT patients to provide an early effective signal of BOS.

**Highlights:** The evolution of peripheral airway function abnormality assessed using Multiple Breath Washout (MBW) and oscillometry following allogeneic HSCT is unknown.

Progressive abnormality is established early following HSCT and occurred in those who developed chronic graft versus host disease (cGHVD) in other organ systems.

This highlights the risk of peripheral airway dysfunction in those affected by cGVHD.

MBW to monitor post-HSCT subjects provides additional insight to that provided by BOS-0p criteria which did not show the same relationship to cGHVD.

These findings identify a potential window for earlier targeted treatment to improve long term outcomes.

## Introduction

Pulmonary complications following Allogeneic Haematopoietic Stem Cell Transplantation (HSCT) are a significant cause of ongoing morbidity and mortality(1). Historically, the only diagnostic pulmonary manifestation of chronic GVHD was biopsy-proven bronchiolitis obliterans, however, its invasive nature and associated risks led to endorsement of adopting the Bronchiolitis Obliterans Syndrome (BOS) approach based on conventional lung function monitoring(2, 3). The incidence of Bronchiolitis Obliterans Syndrome (BOS) reaches 10% in some cohorts(4), but it remains a significant challenge to treat, with overall 2- and 5-year survival of 60% and 50%, respectively(5–8). Poor treatment outcomes may reflect the advanced nature of histological changes at diagnosis. Onset and progression of BOS are attributed to several pre- and post-transplant factors: pre-transplant conditioning regimen, HLA compatibility, respiratory infections and graft versus host disease (GVHD)(9). BOS diagnostic criteria are based on changes in spirometric parameters (FEV_1_ decline of ≥20%) in the setting of an obstructive FEV_1_/FVC ratio(10), but concerns that disease was already advanced by the time of diagnosis led to an additional at-risk category was introduced, BOS-0p or ‘potential BOS’, defined as persistent decline in baseline FEV_1_ of ≥10% and/or baseline FEF_25-75_ of ≥25%(11). However, despite a two year incidence of BOS-0p of 16%(12), the positive predictive value of BOS-0p for BOS was only 29% questioning its utility as an early prediction tool for BOS(12).The limitations of spirometry to detect early changes arising within the peripheral airways has been well documented(13). The peripheral airways contribute relatively little to overall lung resistance, which is a strong determinant of flow in airways (assessed by spirometry). In addition, obstructive lung disease in patchy in nature and adjacent unaffected areas may compensate for any decrease in flow occurring in those affected areas(14). In a study examining the role of lung histology in predicting response to treatment, the authors described better recovery of lung function when histology is consistent with early disease(15). Taken together this highlights the importance of exploring other measures to optimise early disease detection.

Utility of more sensitive peripheral airway function tools, such as Multiple Breath Washout (MBW) and oscillometry, remains under-explored. There is some evidence from cross-sectional studies. In our previous adult study, both MBW parameters (S_acin_ and LCI) and oscillometry parameter reactance (Xrs) were associated with time post-HSCT(16). The relationship with S_acin_ remained significant when adjusted for confounders, was independent of spirometric signs of airflow obstruction or respiratory symptoms(16) and correlated with cGVHD grading. More abnormal S_acin_ and LCI values, compared to spirometry, occurred in histologically confirmed bronchiolitis obliterans (the pathological abnormality in BOS), that correlated with cGVHD grade. In a recent paediatric study of children diagnosed based on symptoms and CT findings, LCI performed far better than FEV_1_ at identifying those affected(17). Collectively, these cross-sectional data suggest the following: that peripheral airway function may worsen over time post-HSCT, this early deterioration is missed by current recommended spirometric monitoring, and that it may relate to the underlying cGVHD process. There is therefore an urgent need to confirm this longitudinally and determine whether worsening of peripheral airway function is driven by evolving BOS (using BOS-0p as its surrogate), cGVHD or both.

Aims of the current longitudinal study were to determine: (1) changes in MBW, oscillometry and spirometric parameters over time, from baseline over 36 months following HSCT; (2) associations of changes with BOS-0p and cGVHD status and severity; and (3) if early MBW and oscillometry changes predicted subsequent BOS-0p development. Hypotheses tested were that significant changes in MBW and oscillometry parameters over time would occur prior to any significant changes in spirometry parameters, and that changes would relate to BOS-0p status and cGVHD severity and predict later BOS-0p status.

## Methods

Longitudinal observational study of allogeneic HSCT recipients at Royal North Shore Hospital, Sydney. All participants underwent (in order) spirometry, plethysmography, gas transfer capacity (DLCO), MBW and oscillometry. Tests were performed at baseline (i.e. pre-HSCT) and 3-monthly intervals thereafter over a 36-month period. Decline from baseline in FEV_1_ of ≥10% led to monthly testing, to ensure any subsequent functional changes were closely monitored. Only data from clinically stable participants free from respiratory infection at time of testing were included in the final dataset. Informed consent occurred at entry to the study which was approved by Northern Sydney Local Health District Human Research Ethics Committee (Protocol Number: LNR/13/HAWKE/11).

### Lung Function testing

Spirometry, plethysmography and DLCO were performed using a V-Max Autobox 6200 plethysmographic system (Sensormedics Co., Yorba Linda, CA, USA), according to American Thoracic Society guidelines(18). Reference values were determined from Global Lungs Initiative equations(19) for spirometry, and Crapo et al. for lung volumes(20) and DLCO(21). Nitrogen (N_2_)-based MBW was performed in triplicate using an in-house N_2_ analyser-based MBW system, to deliver 100% O_2_ during inspiration, as previously described(22, 23). In line with consensus recommendations, participants breathed 100% O_2_ at a tidal volume of 1-1.3 L and a breathing rate of 9-12/min until mean expired N_2_ concentration dropped to <2%(24), reporting mean LCI and FRC from acceptable trials, and S_acin_ and S_cond_ from pooled breath-by-breath Sn_III_ values across three trials(22). MBW parameter z-scores were calculated using published in-house device-specific reference data(25). Oscillometry was performed on an in-house developed device(26) at 6 Hz. Mean respiratory system resistance (Rrs) and reactance (Xrs) were reported across at least three technically acceptable trials, each of 60s duration, as per guideline recommendations(27, 28). Oscillometry parameter z-scores were calculated using published in-house device-specific reference data(26).

### Clinical Data

Clinical data, including information regarding the subject’s diagnosis, donor HLA status, conditioning regimen, acute GVHD (aGVHD) and cGVHD status and grading, were obtained from medical records. Diagnosis and grading of cGVHD was carried out by a senior haematologist as per NIH consensus criteria for grading global cGVHD severity (OLS Table E1)(10). Diagnosis and grading of BOS severity was based on NIH consensus criteria(10) (OLS Table E2). The St George’s Respiratory Questionnaire (SGRQ), completed at each timepoint(29), generated an overall SGRQ score, across symptoms, activity and impact category scores.

### Statistical analyses

Statistical analysis was performed using SPSS (SPSS Inc., Chicago, USA, Version 24). All variables were tested for normality and Z-score transformation was used, where appropriate, to allow comparisons between measured parameters. Statistical significance was defined as p<0.05. Mixed effects modelling examined changes in Z-score transformed spirometry, oscillometry and MBW parameters, vs. days post-HSCT, in a pooled analysis, and separate analyses within groups stratified by BOS and cGVHD status. Changes in measured parameters with days post-HSCT were examined at 3 monthly intervals throughout the 3-year follow up period, to track progression of any statistically significant changes in these parameters. Welch ANOVA tests compared absolute Z-score change in measured parameters between groups. Subjects were stratified by cGVHD status (i.e. cGVHD vs. no cGVHD), as well as global cGVHD grade (1=mild, 2=moderate, 3=severe), for two separate analyses: positive cGVHD status defined as cGVHD at any point after three months post-HSCT, and cGVHD severity defined as maximum global cGVHD grade during follow-up. BOS status was stratified in a similar manner using BOS-0p status given the low number of participants BOS≥1. Smoking history was included as a potential confounding factor.

Logistic regression analyses examined associations between early MBW changes at 90 or 180 days post-HSCT and development of BOS-0p at 1, 2 and 3 years. Given the lack of known thresholds for clinically meaningful change, utility of different absolute Z-score change were explored in baseline steps from 0.5 to 2 Z-scores (using 0.5 steps). Only peripheral airway function parameters that changed significantly on mixed modelling analysis at time points 90 or 180 days post-HSCT were included in logistic regression analyses. One way ANOVA with Tukey’s HSD post-hoc test was performed to compare absolute Z-score change in MBW and oscillometry parameters between groups stratified by NIH cGVHD severity grade (0, 1, 2 and 3). Correlations between change in MBW and oscillometry parameters and other clinical parameters were examined using Pearson’s or Spearman’s correlation analyses. Changes in spirometry, MBW or oscillometry parameters were defined as absolute Z-score change from baseline, measured at the subject’s final follow-up visit. Receiver Operator Characteristic (ROC) analyses were performed comparing MBW and Spirometry parameters as predictors for cGVHD, BOS-0p and BOS at 6 months and 12 months post-HSCT.

## Results

69 allogeneic HSCT recipients were recruited, of which 64 were included in the analysis: five participants were excluded due to development of purely restrictive lung disease. Clinical, anthropometric and baseline pulmonary and peripheral airway function data of included participants are summarised in Tables 1 and 2, respectively. The vast majority had an HLA-matched donor and a myeloablative conditioning regimen. The proportion of the cohort followed up at 1 year, 2 years and 3 years was 51/64 (80%), 38/64 (59%) and 28/64 (44%), respectively. Clinical and baseline characteristics of those participants lost to follow up did not differ from those retained within the study (OLS Table E4), apart from a higher proportion of males (67%, p=0.013) and higher baseline S_acin_ values (mean difference 0.03 L^-1^, p=0.014).

**Table 1.** Clinical characteristics of HSCT recipients at baseline across the entire cohort, within groups stratified by BOS-0p and cGVHD status.

|  | *All Subjects* | *BOS-0p* | *No BOS-0p* | *cGVHD* | *No cGVHD* |
| --- | --- | --- | --- | --- | --- |
| **N (males, %)** | 64 (52%) | 23 (50%) | 41 (56%) | 46 (63) | 18 (26)** |
| **Age (years)** | 50 ± 12 (18-68) | 52 ± 11 (21-68) | 50 ± 13 (18-68) | 51 ± 13 (18-68) | 51 ± 11 (25-66) |
| **Height (cm)** | 171 ± 9 | 172 ± 8 | 171 ± 10 | 173 ± 9 | 166 ± 7** |
| **BMI (kg/m<sup>2</sup>)** | 20 ± 4 | 27 ± 5 | 26 ± 4 | 26 ± 4 | 26 ± 6 |
| **Smoking History** | 6 ± 10 (0-50) | 3 ± 6 (0-20) | 8 ± 12 (0-50) | 6 ± 11 (0-50) | 5 ± 8 (0-20) |
| **Total Follow Up Period (days)** | 715 ± 430 | 772 ± 369 | 682 ± 462 | 705 ± 444 | 772 ± 376 |
| **Diagnosis** |  |  |  |  |  |
| AML | 20 (31) | 6 (26) | 14 (34) | 17 (37) | 3 (17) |
| ALL | 10 (16) | 5 (22) | 5 (12) | 6 (13) | 4 (22) |
| LPD | 20 (31) | 8 (35) | 12 (30) | 16 (35) | 4 (22) |
| Myeloma | 5 (8) | 2 (9) | 3 (7) | 3 (7) | 2 (11) |
| MDS | 9 (14) | 2 (9) | 7 (17) | 4 (8) | 5 (28)* |
| **Donor HLA Status** |  |  |  |  |  |
| HLA-matched related | 44 (69) | 17 (74) | 27 (66) | 35 (76) | 9 (50) |
| HLA-matched unrelated | 15 (23) | 5 (22) | 10 (24) | 9 (20) | 6 (33) |
| HLA-mismatched unrelated | 5 (8) | 1 (4) | 4 (10) | 2 (4) | 3 (17) |
| **Conditioning Regimen** |  |  |  |  |  |
| Myeloablative | 45 (70) | 16 (70) | 29 (70) | 31 (67) | 12 (67) |
| Reduced Intensity | 19 (30) | 7 (30) | 12 (30) | 15 (33) | 6 (33) |
| Total Body Irradiation | 29 (45) | 14 (58) | 15 (37) | 21 (46) | 8 (44) |

Footnote: Data displayed as Mean±SD or N (%) unless otherwise stated. AML, acute myeloid leukaemia; ALL, acute lymphoblastic leukaemia; BOS, bronchiolitis obliterans syndrome; cGVHD, chronic graft versus host disease; LPD, lymphoproliferative disease; MDS, Myelodysplastic syndrome; HLA, human leukocyte antigen. Asterisks indicate a statistically significant difference, in either the mean or proportion (%) of clinical characteristics, between BOS-0p and No BOS-0p groups, or cGVHD and No cGVHD, respectively, at the following levels: \*p<0.05, \*\*p<0.01, \*\*\*p<0.001.

Twenty-three (36%) participants fulfilled BOS-0p criteria during follow up with a mean±SD time to onset of 411±269 days post-HSCT. Baseline FEV_1_ %predicted (p=0.001) and FEF_25-75_ %predicted (p=0.007) values were significantly higher in BOS-0p vs. No BOS-0p participants, whilst No BOS-0p participants had significantly higher RV/TLC values (p=0.002, Table 2). Of the 23 BOS-0p participants, four (17%) later developed NIH-defined BOS; two patients with BOS stage 1, one with BOS stage 2 and one with BOS stage 3. Thirty-two participants (50%) developed aGVHD (OLS Table E3), and 46 (72%) cGVHD during follow up (onset 266±198 days). The proportion developing cGVHD was comparable across those with and without BOS-0p: 74% vs. 68% (p=0.62), respectively.

**Table 2.** Baseline Pulmonary and Peripheral Airway Function across the entire cohort and within groups stratified by BOS-0p and cGVHD status.

|  | *All Subjects* | *BOS-0p* | *No BOS-0p* | *cGVHD* | *No cGVHD* |
| --- | --- | --- | --- | --- | --- |
| **Pulmonary Function** |  |  |  |  |  |
| FEV<sub>1</sub> %pred | 102 ± 15 | 110 ± 12 | 98 ± 15** | 103 ± 13 | 100 ± 19 |
| FEV<sub>1</sub>/FVC (%) | 78 ± 6 (64-90) | 79 ± 6 (67-90) | 77 ± 6 (64-90) | 78 ± 6 (64-90) | 78 ± 7 (70-90) |
| FEF<sub>25-75</sub> (%) | 83 ± 28 | 96 ± 26 | 76 ± 27** | 84 ± 23 | 83 ± 40 |
| TLC %pred | 99 ± 11 | 98 ± 11 | 99 ± 11 | 99 ± 11 | 98 ± 12 |
| RV/TLC (%) | 31 ± 7.5 | 27 ± 6.2 | 33 ± 7.3** | 30 ± 7 | 32 ± 9 |
| DLCO %pred | 80 ± 14 | 84 ± 15 | 78 ± 12 | 80 ± 14 | 79 ± 14 |
| Lung Function Score | 2.9 ± 0.93 | 2.6 ± | 3.1 ± 0.97 | 2.8 ± 0.90 | 3.8 ± 0.99 |
| **SGRQ Total Score** | 13 ± 11 | 13 ± 10 | 12 ± 11 | 11 ± 9 | 16 ± 13 |
| **Peripheral Airway Function** |  |  |  |  |  |
| S<sub>acin</sub> (L<sup>-1</sup>) | 0.10 ± 0.05 | 0.096 ± 0.04 | 0.103 ± 0.05 | 0.101 ± 0.05 | 0.100 ± 0.04 |
| S<sub>cond</sub> (L<sup>-1</sup>) | 0.03 ± 0.01 | 0.029 ± 0.01 | 0.033 ± 0.02 | 0.03 ± 0.01 | 0.034 ± 0.02 |
| LCI (lung turnovers) | 8.58 ± 1.10 | 8.30 ± 0.94 | 8.74 ± 1.12 | 8.33 ± 0.91 | 9.16 ± 1.31 |
| Rrs (cmH<sub>2</sub>O.L<sup>-1</sup>.s<sup>-1</sup>) | 3.57 ± 1.29 | 3.58 ± 1.51 | 3.57 ± 1.14 | 3.31 ± 1.13 | 4.24 ± 1.43 |
| Xrs (cmH<sub>2</sub>O.L<sup>-1</sup>.s<sup>-1</sup>) | -0.66 ± 0.5 | -0.60 ± 0.46 | -0.69 ± 0.53 | -0.60 ± 0.55 | -0.81 ± 0.34 |

Footnote: Data displayed as Mean±SD or N (%) unless otherwise stated. BOS, bronchiolitis obliterans syndrome; cGVHD, chronic graft versus host disease. Asterisks indicate a statistically significant difference, in either the mean or proportion (%) of pulmonary and peripheral function parameters, between BOS-0p and No BOS-0p groups, or cGVHD and No cGVHD, respectively, at the following levels: \*p<0.05, \*\*p<0.01, \*\*\*p<0.001.

### Changes in lung function parameters over time

Changes in MBW, oscillometry and spirometric parameters are summarised in Figure 1 (as absolute change from baseline at each time point) and in Table 3 (as rate of z-score change per month from baseline at each time point) for the first year. Corresponding values for the entire three year follow up are shown in OLS Figure E1 and Table E5. 51/64 (81%) participants were followed up at one or more time points within the first year post-HSCT (Table 3). Across the entire cohort, S_acin_ and LCI were the first lung function parameters to significantly change, as early as 90 days post-HSCT, before any other measured parameter: S_acin_ 0.224 (p<0.001) and LCI 0.126 (p=0.03), respectively, data displayed as mixed model estimate [rate of change in Z-score per month]. Greatest rate of increase in S_acin_ and LCI z-scores occurred within the first 180 days (OLS Table E5). Statistically significant changes in other measured parameters were observed at later time points, in the following order: FEF_25-75_ at 450 days (−0.017, p=0.04), S_cond_ (0.032, p=0.002) and FEV_1_ /FVC (−0.016, p=0.007) at 630 days, and Xrs at 720 days post-HSCT (−0.039, p=0.03). By contrast, change in Rrs was close to, but did not reach, statistical significance at 1080 days (0.021, p=0.06).

**Figure 1.**
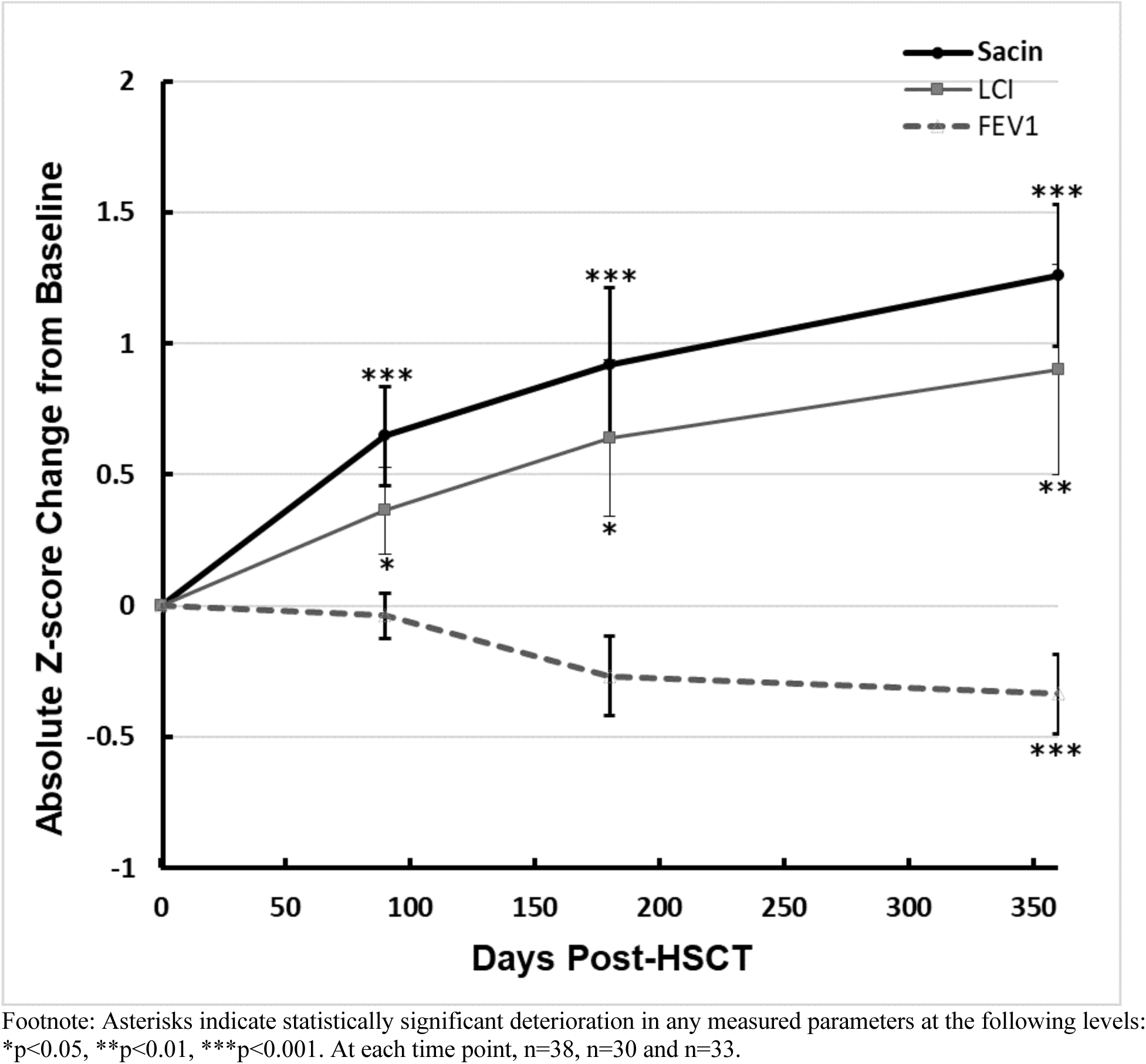
Change in S_acin_, LCI and FEV_1_ from baseline within the first year post-HSCT, across the entire cohort.

**Table 3.** Estimated rates of Z-score change per month in S_acin_, LCI and FEV_1_ within the first year post-HSCT, across the entire cohort.

| Days Post HSCT | Parameter | *All Subjects* |  |  |
| --- | --- | --- | --- | --- |
|  |  | estimate* | S.E. | p value |
| **90**<br>n=38 | S<sub>acin</sub> Z-score | 0.20 | 0.05 | <0.001 |
|  | LCI Z-score | 0.12 | 0.05 | 0.04 |
|  | FEV<sub>1</sub> Z-score | -0.01 | 0.02 | 0.54 |
| **180**<br>n=30 | S<sub>acin</sub> Z-score | 0.17 | 0.03 | <0.001 |
|  | LCI Z-score | 0.09 | 0.04 | 0.03 |
|  | FEV<sub>1</sub> Z-score | -0.03 | 0.02 | 0.09 |
| **360**<br>n=33 | S<sub>acin</sub> Z-score | 0.11 | 0.01 | <0.001 |
|  | LCI Z-score | 0.06 | 0.02 | <0.01 |
|  | FEV<sub>1</sub> Z-score | -0.03 | 0.01 | <0.001 |

Footnote: \*Mixed model estimates represent the rate of change in Z-score per month

### Changes in lung function parameters stratified by BOS-0p status

In both BOS-0p and No BOS-0p groups, S_acin_ was also the first parameter to change, at 90 days post-HSCT, before any significant changes in any other measured parameter (Figure 2, OLS Table E6 and E7). In the BOS-0p group both FEV_1_ and FEF_25-75_ declined at 180 and 360 days, respectively. Statistically significant deteriorations in other parameters occurred in the following order: LCI at 180 days (0.13, p=0.037), FEV_1_/FVC (−0.044, p=0.004) and Xrs at 360 days (−0.107, p=0.03) and S_cond_ at 540 days post-HSCT (0.05, p=0.007). In the No BOS-0p group, there were no significant deteriorations in spirometry or other measured parameters within the first year, or at any timepoint examined. While overall change in S_acin_ was numerically greater in the BOS-0p group, vs. No BOS-0p, this between-group difference did not reach statistical significance within the first year [90 (p=0.515), 180 (p=0.703), or 360 days (p=0.303)] or at any later timepoints.

**Figure 2.**
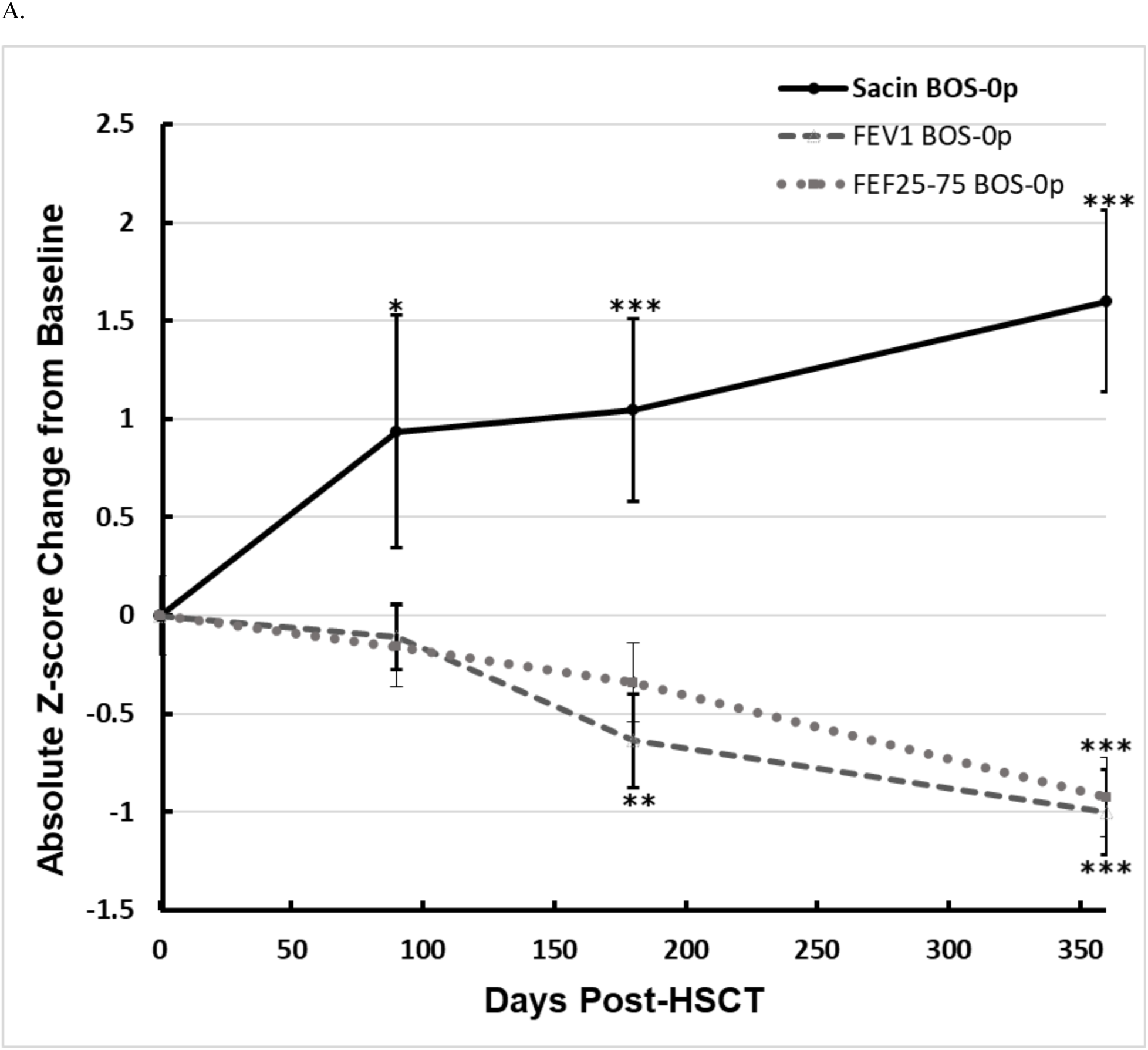
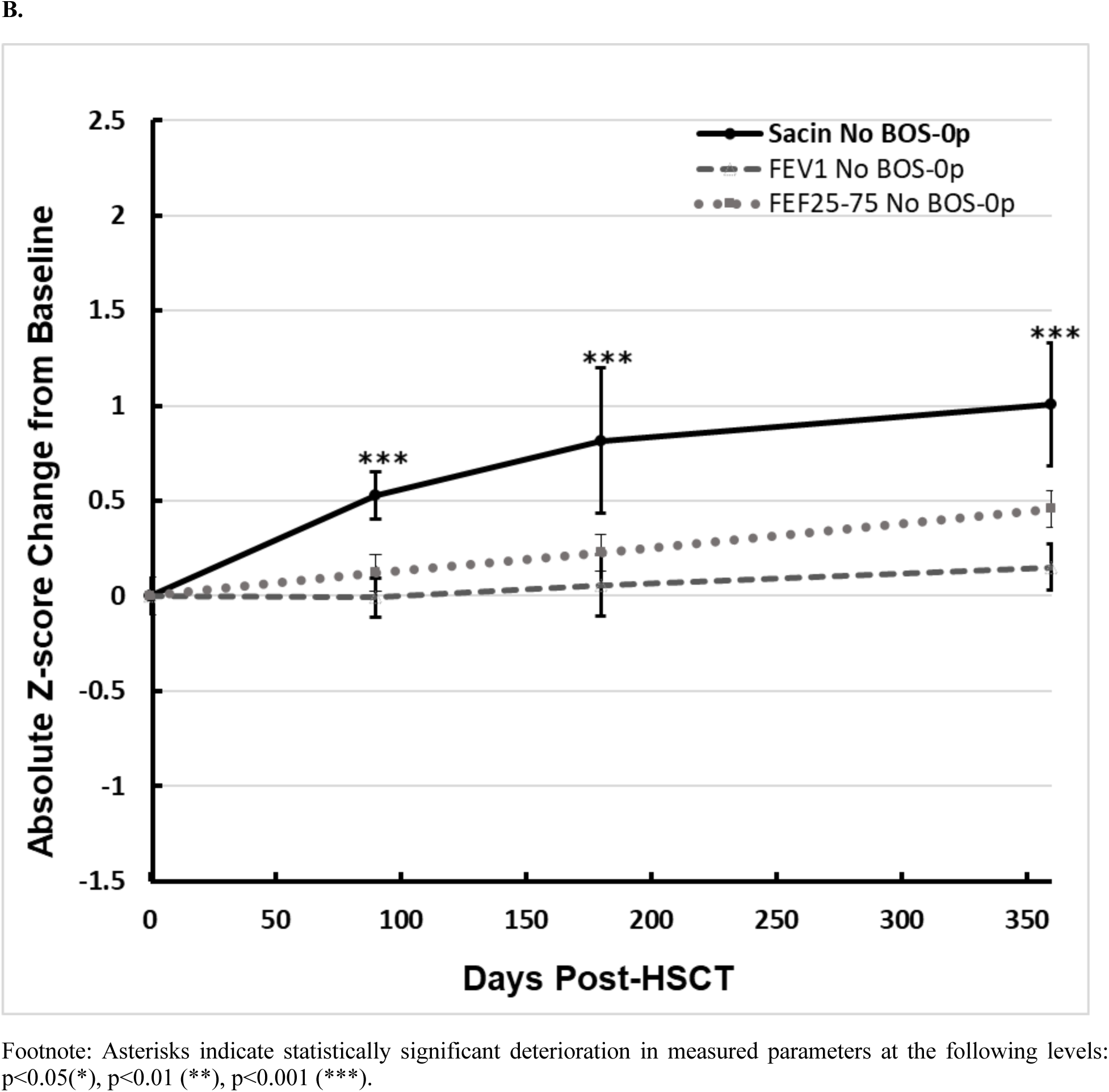
Change in S_acin_, FEV_1_ and FEF_25-75_ within the first year post-HSCT, in BOS-0p (A) and No BOS-0p (B) groups.

### Changes in lung function parameters stratified by cGVHD status and severity

When comparing groups stratified by cGVHD status (Figure 3, OLS Tables E8 and E9), S_acin_ and LCI were the first parameters to change (in Z-score value) at 90 days, whilst significant changes in any other measured parameter were not detectable until one year: S_acin_ 0.25 (p=0.001) and LCI 0.21 (p=0.01) in the cGVHD group. The greatest rate of increase in S_acin_ and LCI z-scores occurred within the first 90 days. Statistically significant deteriorations in other measured parameters were observed in the cGVHD group at later time points, beyond the first year, in the following order: FEV_1_ at 360 days (−0.044, p<0.001), FEF_25-75_ (−0.022, p=0.04) and Xrs (−0.058, p=0.048) at 450 days, S_cond_ at 540 days (0.036, p=0.03), FEV_1_ /FVC at 630 days (−0.021, p=0.01), and Rrs at 1080 days post-HSCT (0.023, p=0.03). In the No cGVHD group, time to significant deterioration, if it occurred, was much longer for all parameters examined, with spirometry and oscillometry parameters all unchanged throughout follow up, S_acin_ only transiently so between 540 and 630 days post-HSCT, and S_cond_ almost reaching statistical significance at 900 days post-HSCT (0.029, p=0.052). Between-group difference in S_acin_ reached statistical significance from 180 days post-HSCT onwards (OLS Table E10), as it did for LCI, although for LCI the difference was not maintained at one year (OLS Table E11, p=0.065). No change was seen for FEV_1_ (OLS Table E12).

**Figure 3.**
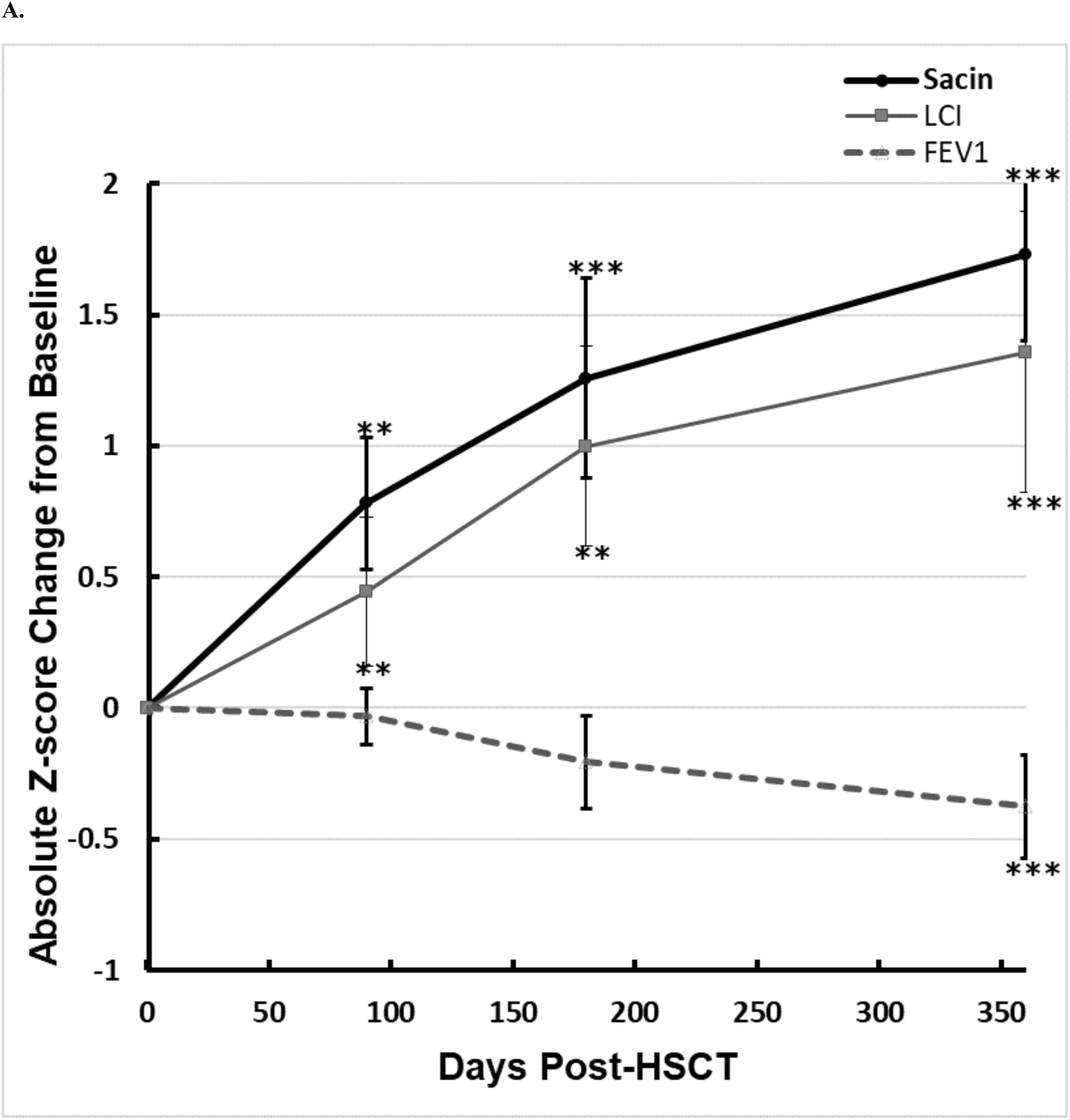
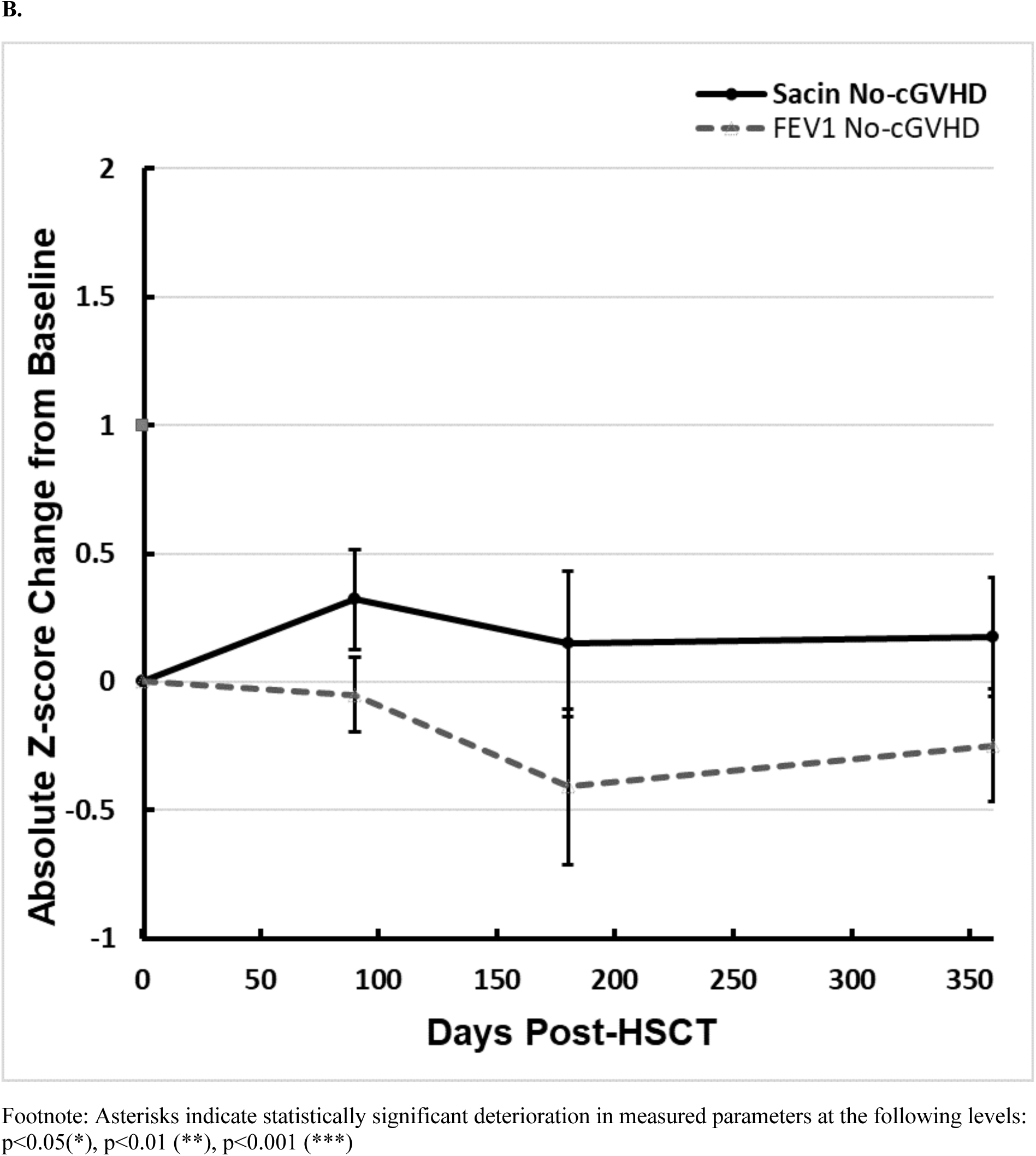
Change in S_acin_, LCI and FEV_1_ with number of days post-HSCT in cGVHD (A) and No cGVHD (B) groups.

Absolute Z-score change from baseline in S_acin_ was related to the severity of cGVHD severity grade (OLS Table E13): cGVHD grade 3 had significantly greater worsening of S_acin_ during follow up, compared to cGVHD grades 0, 1 and 2. S_acin_ changes in grades 0, 1 and 2 were similar. Z-score changes in LCI and FEV_1_ between cGVHD grades were similar (data not shown).

### Ability of lung function parameters to predict BOS-0p and cGVHD outcomes

Changes in S_acin_ were not predictive for later BOS-0p development on logistic regression analyses, although associations between change in S_acin_ Z-score thresholds ≥1 and ≥1.5 at 180 days and BOS-0p 1 and 2 years approached statistical significance (p=0.07) (OLS Table E14). Similarly, baseline Z-score values for S_acin_ and LCI were not predictive of later BOS-0p, nor were changes in LCI Z-score at 90 or 180 days. Similarly, ROC analysis showed no predictive value of MBW parameters for BOS-0p status (OLS Table E15). ROC analysis showed that whilst some MBW parameters were predictive of cGVHD – LCI at 6 months for later cGVHD and S_acin_ at 12 months for current or later cGVHD – a number of participants developed cGVHD (of other organs systems) either before or after the measurement time point, such that MBW parameters were not providing a consistent early signal (OLS Table E16).

## Discussion

In summary, this longitudinal observation study of lung function changes post allogeneic HSCT outlined that increases in S_acin_ were common post-HSCT and occurred early, within the first six months. Changes in Xrs and spirometry occurred but later, while Rrs did not change at all during the three-year follow up period. S_acin_ change was associated with cGVHD; whilst it increased progressively from 90 days onwards in those with cGVHD, spirometry parameters only changed nine months later. However, no changes to any peripheral airway parameters were associated with BOS-0p, compared to those without BOS-0p, despite BOS-0p occurring commonly in this group, and S_acin_ changes occurred independently. BOS 1 or greater was uncommon, affecting only four participants, each of which had earlier BOS-0p and S_acin_ abnormality. Therefore early changes in S_acin_ (or any other lung function parameter) were not predictive of BOS-0p, and the association with cGVHD is consistent with lung involvement in the cGVHD process, and was not visible by spirometry. Taken together, these findings suggest that MBW provides differing insight to BOS-0p criteria, and because of its strong association with cGVHD may be of greater clinical relevance, a signal missed by the currently proposed spirometry-based at risk criteria.

These findings provide novel longitudinal evidence of the potential value of MBW for detecting early peripheral airway dysfunction within a large adult cohort of HSCT survivors. Deterioration in acinar ventilation inhomogeneity (S_acin_) occurred early, by 90 days, and prior to any significant change in spirometry or oscillometry parameters. S_acin_ was not significantly greater in participants with BOS-0p, compared to those without BOS-0p, nor did early deterioration in S_acin_ predict later BOS-0p. However, S_acin_ was significantly greater in participants with cGVHD, compared to those without cGVHD, and rate of deterioration in S_acin_ was related to cGVHD severity. This strong association between S_acin_, LCI and cGVHD confirms prominent lung involvement in the cGVHD process. Importantly, there does not appear to be a pre-determined order in which organs become involved in the cGVHD process, and our data suggests that whilst abnormalities in MBW S_acin_ and LCI parameters are associated with cGVHD, abnormality does not consistently predict later cGVHD as the lung may not always be the first organ involved.

Across the entire cohort, change in S_acin_ was accompanied by significant worsening of global ventilation heterogeneity (LCI). By contrast, significant change in S_cond_ and Xrs occurred much later, at 630 and 720 days respectively, and Rrs showed no significant change over time post-HSCT. These results confirm the previous cross-sectional findings of Lahzami et al(16) of an association between both S_acin_ and LCI and time post-HSCT, independent of spirometric signs of airflow obstruction or respiratory symptoms. Additionally, within our longitudinal cohort S_acin_ deterioration occurred much earlier than S_cond_ suggesting that early peripheral airway dysfunction post-HSCT may predominantly arise in more distal airway architecture within the region of the acinar entrance, with more proximal conducting airways affected at a later stage. These findings are largely consistent with histological findings in bronchiolitis obliterans, where terminal and respiratory bronchioles were predominantly affected(30, 31).

As the current proposed functional screening tool for evolving abnormality seen with bronchiolitis obliterans, BOS-0p status was not a good discriminator of changes in S_acin_. Deteriorations in S_acin_ were not exclusive to those meeting BOS-0p criteria: significant worsening of S_acin_ over time occurred both in those with and without BOS-0p, with no significant difference between groups, and early S_acin_ changes were not predictive of later BOS-0p. Whilst we did not have sufficient numbers of BOS in our study, there is published evidence supporting a predictive role of indices of ventilation distribution for BOS in another setting, post lung transplantation: abnormal phase III slope (S_III_) from N_2_ single breath washout (SBW) preceded BOS1 by 168 days(32). Interaction between gas diffusion and convection predominant in the acinar airways, reflected by S_acin_ in MBW, is recognised to be a significant contributor to S_III_ in SBW(33). We would propose the lack of discrimination within our cohort represents the weakness in the ability of BOS-0p to predict the pathophysiological process underlying BOS, as others have highlighted in both HSCT(12) and post lung transplantation(34). The incidence of BOS-0p was relatively high in our study as it is elsewhere in the literature: 36% within the first three years post HSCT in our cohort and 16% within the first two years in the study by Abedin et al(12). The positive predictive value of BOS-0p for BOS in the Adedin et al cohort was only 29%(12). An additional criterion of reduced FEV_1_/FVC ratio for BOS-0p has been recommended(35) but would have had little impact on early disease detection in our cohort as only 4 of 23 BOS-0p participants (17%) had co-existing reduced FEV_1_/FVC ratio. Finally, cGVHD occurred in an equal proportion of participants in both BOS-0p and No BOS-0p groups in our cohort.

Our data suggests an important role of the lungs in the cGVHD process. We have demonstrated previous cross-sectional associations between S_acin_ and cGVHD(16, 36, 37) but through this longitudinal study, we now show that (i) early change in S_acin_ occurred almost exclusively in HSCT recipients either with or destined to develop cGVHD, before any significant deteriorations in spirometry; and (ii) S_acin_ was significantly and persistently greater in the cGVHD group, compared with the No cGVHD group, from 180 days post-HSCT. Failure to observe the same pattern in LCI (significantly different at 180 days but p=0.065 at 360 days) may reflect a type 2 error. Magnitude of overall deterioration in S_acin_ was determined by severity of cGVHD grade, confirming findings of Nyilas et al(37) and Lahzami et al(16). Whilst this may be expected, given that lung score, defined based on spirometry changes, contributes to cGVHD scores ≥2, it should be noted that this cohort contained only four BOS≥1 participants. Importantly, cGVHD diagnosis using conventional criteria occurred both before and after the development of S_acin_ abnormality, suggesting that MBW-detected abnormalities may at times reperesent the first organ affected by cGVHD, or may be useful as a marker of severity by indicating additional organ involvement. Our results support further studies to evaluate the utility of MBW as a surveillance tool in post-HSCT subjects. The widespread availability of validated commercial devices(38) and consensus MBW guidelines for test performance and data analysis(24) have greatly increased the feasibility of widespread testing. This improves the feasibility of large multicentre studies to determine how early MBW changes relate to later BOS occurrence, a critical step in determining the prognostic utility of the changes we describe here. Interestingly, deterioration in S_acin_ occurred early, at a time when most immunosuppressive therapy is typically weaned post-HSCT. This suggests that should the prognostic utility of MBW to detect BOS be confirmed, subsequent studies could also explore the utility of MBW monitoring to guide future immunosuppression withdrawal, regardless of whether other organ cGVHD is evident, and improve post-HSCT pulmonary outcomes.

There are several strengths to this study. Z-scores for MBW parameters, the focus of our study, were generated using published normative data specific to our device(25). These reference values previously highlighted the significant increase in ventilation inhomogeneity that occurs with age in healthy subjects, and age-specific Z-scores were therefore an important strength in our current longitudinal study of subjects across a wide age range at HSCT (18-68 years), and for increases in age occurring in subjects over the follow up period. Monitoring was performed at frequent time points post-HSCT, avoiding times of acute intercurrent illness, allowing us to describe accurately the trajectories observed free from the impact of acute infection. Detailed clinical objective assessment of cGVHD status by a haematologist was critical to describing relationships present. However, there are also notable limitations. As a single centre study, we did not have sufficient numbers of participants with proven BOS (i.e. BOS≥1) to investigate and compare MBW and oscillometry utility beyond the BOS-0p setting. This will be best accomplished by future multi-centre studies. The limited prognostic utility of BOS-0p to detect BOS has been described, with only 29% of BOS-0p subjects progressing to BOS(12). The correlation of MBW with cGVHD, but not BOS-0p, suggests that MBW is providing additional insight to BOS-0p and may be of greater clinical relevance, given cGVHD is a well-established risk factor for BOS(1). Whilst our study numbers were sufficient to demonstrate difference in rate of S_acin_ deterioration between those with and without cGVHD, and between severe (grade 3) and lesser severity cGVHD (grade 0-2), they were insufficient to explore relationships between individual grades. We also lack histological evidence of lung GVHD in our study, a common issue in studies due to the decreasing use of lung biopsy in this setting, of concurrent HRCT data to look for evidence of gas trapping, which would be supportive of pulmonary GVHD. In our analysis, we also had to adjust for variable adherence to our follow-up schedule of 3-monthly visits. Additionally, whilst retention of participants within the longitudinal cohort was challenging, with only 28/64 participants (44%) completing follow up to three years, the vast majority 51/64 (80%) were retained for follow up to one year and numbers were still sufficient to demonstrate statistically significant early changes in peripheral airway function, as well as strong associations between these changes and cGVHD. Clinical and baseline functional characteristics of those retained throughout the study were comparable to those lost to follow up, suggesting strong representation of the overall cohort.

In summary, this study provides novel longitudinal evidence of early progressive peripheral airway dysfunction, detected by MBW, after allogeneic HSCT. Importantly, S_acin_ abnormality was the earliest detectable change in this setting, prior to any change in spirometry parameters. cGVHD status, and not BOS-0p, was strongly associated ith these longitudinal deteriorations in peripheral airway function, independent of spirometry changes (and BOS-0p status). These findings suggest an important role of the lungs in the cGVHD process, which may be the initial organ affected. We feel that our findings question the utility of BOS-0p as an at risk stage for pulmonary GVHD, and provide important evidence to support future studies to investigate the clinical relevance of deteriorations in S_acin_ after HSCT.

## Contributors

Study Design: all authors. Recruitment, patient testing and data collection: CH, RS, MG, VP, SH. Statistical analysis CH, AP, JH and CT. Manuscript writing: all authors.

## Data Availability

All data produced in the present study are available upon reasonable request to the authors

