## Supplementary material for "Early Progressive Peripheral Airway Dysfunction after Allogeneic Haematopoietic Stem Cell Transplantation is associated with chronic Graft vs Host disease not BOS-0p": Online supplement

### 1. Definitions used

#### 1.1 Definition of chronic GVHD (cGVHD)

**Table E1**. NIH Criteria for Grading the Global Severity of cGVHD[1]

| NIH Global cGVHD Grade | Criteria |
|---|---|
| **1**<br>**Mild** | 1 or 2 Organs involved with no more than score 1* plus<br>Lung score 0 |
| **2**<br>**Moderate** | 3 or More organs involved with no more than score 1<br>OR<br>At least 1 organ (not lung) with a score of 2<br>OR<br>Lung score 1 |
| **3**<br>**Severe** | At least 1 organ with a score of 3<br>OR<br>Lung score of 2 or 3 |

Footnote: *Further details of the clinical scoring of cGVHD affecting specific organ systems are provided in the 2014 NIH Diagnosis and Staging Working Group Report[1].

#### 1.2 Definition of BOS

Diagnosis of BOS relied heavily on spirometric evidence of new onset fixed airflow obstruction, in the absence of acute respiratory infection. In the presence of another distinctive manifestation of cGVHD, BOS was considered a physiological surrogate for pulmonary GVHD, provided that the following criteria were met: an $FEV_1/FVC$ ratio <0.7 or <LLN, an $FEV_1$<75% predicted with a ≥10% decline within a 2 year period, as well as some features such as air trapping on PFT's (RV/TLC >ULN) or expiratory CT [1]. Additionally, the criteria added that bronchial biopsy would only be required to confirm the diagnosis of pulmonary GVHD, if BOS criteria were satisfied without any other distinctive manifestation of cGVHD. In the few HSCT survivors who developed BOS, during their follow-up period, the severity of BOS was further classified according to the extent of decline in baseline FEV1, as per International Society for Heart and Lung Transplantation (ISLHT) guidelines; this is summarized in Table E2.

**Table E2**. ISLHT Criteria for Grading the Global Severity of cGVHD [1,2].

| BOS Stage | Criteria |
|---|---|
| **BOS 0-p** | decline in baseline $FEV_1$ ≥10% and or baseline $FEF_{25-75}$ ≥25% |
| **0** | $FEV_1$: ≥80% of baseline |
| **1** | $FEV_1$: 66-80% of baseline |
| **2** | $FEV_1$: 51-65% of baseline |
| **3** | $FEV_1$: ≤50% of baseline |

Due to an insufficient number of patients with BOS in our HSCT cohort, subjects were stratified by BOS stage 0-p (BOS 0-p) status, according to the criteria proposed by Estenne et al. (2001)[2] for identifying those at risk of developing BOS. BOS 0-p or '*potential BOS*' was defined as a persistent decline in baseline $FEV_1$ ≥10% and or baseline $FEF_{25-75}$ ≥25%. Some studies have shown the prognostic value of adding an obstructed $FEV_1/FVC$ ratio of <80% predicted to the above BOS 0-p criteria [3]. Although this could potentially improve the sensitivity for detecting BOS Stage 1 from 57% using the BOS 0-p criteria alone to 71%, our classification of subjects was based purely on the persistent decline in $FEV_1$ and $FEF_{25-75}$.

**1.3 Definition of severity of acute and chronic GVHD**

Acute GVHD (aGVHD) and cGVHD severity grading for the cohort are summarised in Table E3, for all subjects and within groups stratified by BOS-0p and cGVHD status. There were a proportionately greater number of subjects with lung involvement in the cGVHD grade 3 group, as compared with cGVHD grade 2 (p=0.01), and cGVHD grade 1 (p<0.001). Of the 4/7 (57%) patients in the cGVHD grade 3 group with lung involvement, two had NIH defined BOS (stages 2 and 3) and two went onto developing BOS stage 3, soon after their 2nd HSCT. For the latter two patients, we only included data preceding their 2nd HSCT in the analysis. However, within this follow-up period, both these subjects had experienced deteriorations in $S_{acin}$ of up to an absolute Z-score change of 3.7 SD from baseline, in the absence of any deterioration in $FEV_1$. By contrast, there were only 2/20 (10%) subjects with less severe BOS stage 1 in the cGVHD grade 2 group, and none with NIH defined lung involvement in the cGVHD grade 1 group.

**Table E3**. aGVHD and cGVHD severity grades across the entire cohort and within groups stratified by BOS-0p and cGVHD status.

| | *All Subjects* | *BOS-0p* | *No BOS-0p* | *cGVHD* | *No cGVHD* |
|---|---|---|---|---|---|
| **aGVHD Grade** | | | | | |
| 0 | 32 (50) | 11 (48) | 21 (51) | 21 (46) | 11 (61) |
| 1 | 11 (17) | 3 (13) | 8 (20) | 9 (20) | 2 (11) |
| 2 | 19 (30) | 8 (35) | 11 (27) | 14 (30) | 5 (28) |
| 3 | 1 (2) | 1 (4) | 0 | 1 (2) | 0 |
| 4 | 1 (2) | 0 | 1 (2) | 1 (2) | 0 |
| **cGVHD Grade** | | | | | |
| 0 | 18 (28) | 6 (26) | 13 (32) | -- | 18 (100) |
| 1 | 19 (30) | 5 (22) | 13 (32) | 19 (41) | -- |
| 2 | 20 (31) | 9 (39) | 11 (27) | 20 (44) | -- |
| 3 | 7 (11) | 3 (13) | 4 (10) | 7 (15) | -- |
| **cGVHD Onset** | 266 ± 198 | 250 ± 175 | 276 ± 216 | 266 ± 198 | -- |
| **BOS-0p Onset** | -- | 411 ± 269 | -- | -- | -- |

Footnote: Data displayed as Mean±SD or N (%) unless otherwise stated. Note that the aGVHD (Glucksberg Criteria) and cGVHD (NIH Consensus Criteria) grades reported are the maximum grades reached during follow-up. Asterisks or ϕ indicate a statistically significant difference, in either the mean or proportion (%) of clinical characteristics, between BOS-0p and No BOS-0p groups, or cGVHD and No cGVHD, respectively, at the following levels: *p<0.05, **p<0.01, ***p<0.001.

#### 2. Characteristics of subjects lost to follow up

When comparing the 36 patients lost to follow up (LTFU) to the cohort that were retained in the study to complete their study visits, the vast majority of clinical and baseline functional characteristics were comparable (Table E4); the proportion of males was significantly higher (67%, p=0.013) in the LTFU group; similarly, baseline $S_{acin}$ values were also higher in this group (mean difference = 0.03, p=0.014). Thirty of the 36 (83%) who were LTFU showed no signs of respiratory involvement; of the remaining 6 patients, 5 were reported to have experienced respiratory symptoms due to pneumonia, bronchitis, scleroderma, subglottic extrapulmonary plasmocytoma and lomentospora prolificans infection; whereas only 1 patient had respiratory symptoms due to BOS and this patient withdrew from the study at 2 years, having contributed data at that time point. Fifteen of the 18 (83%) deaths in the LTFU group were also due to non-respiratory causes such as transplant related complications and those associated with relapsed haematologic malignancies. The remaining 3 deaths included the patient with BOS, who died 6 years post-HSCT, as well as 1 other who died from disseminated lomentospora prolificans infection following their 2nd HSCT, and 1 patient who died of unknown causes.

**Table E4**. Baseline (a) clinical and (b) functional characteristics across groups stratified by lost to follow up (LTFU) status.

**(a) Clinical Characteristics**

| | **Retained** | **LTFU** |
|---|---|---|
| **N (males, %)** | 28 (36%) | 36 (67%)* |
| **Age (years)** | 50 ± 11 (24-68) | 51 ± 13 (18-68) |
| **Height (cm)** | 169 ± 10 | 172 ± 8 |
| **BMI (kg/m$^2$)** | 26 ± 5 | 26 ± 4 |
| **Smoking History** | 3 ± 8 (0-35) | 8 ± 12 (0-50) |
| **Total Follow Up Period (days)** | 1141 ± 205*** | 400 ± 219 |
| **Diagnosis** | | |
| AML | 11 (40) | 9 (25) |
| ALL | 4 (14) | 6 (17) |
| LPD | 9 (32) | 11 (31) |
| Myeloma | 2 (7) | 3 (8) |
| MDS | 2 (7) | 7 (19) |
| **Donor HLA Status** | | |
| HLA-matched related | 20 (71) | 24 (67) |
| HLA-matched unrelated | 3 (11) | 11 (30) |
| HLA-mismatched unrelated | 5 (18) | 1 (3) |
| **Conditioning Regimen** | | |
| Myeloablative | 11 (39) | 9 (25) |
| Reduced Intensity | 17 (61) | 27 (75) |
| Total Body Irradiation | 12 (43) | 17 (47) |
| **cGVHD Grade** | | |
| 0 | 10 (36) | 9 (25) |
| 1 | 4 (14) | 14 (39) |
| 2 | 11 (39) | 9 (25) |
| 3 | 3 (11) | 4 (11) |

Footnote: Data displayed as Mean±SD or N (%) unless otherwise stated. AML, acute myeloid leukaemia; ALL, acute lymphoblastic leukaemia; cGVHD, chronic graft versus host disease; LPD, lymphoproliferative disease; MDS, Myelodysplastic syndrome; HLA, human leukocyte antigen.

Asterisks indicate a statistically significant difference, in either the mean or proportion (%) of clinical characteristics, between those who completed their study visits (LTFU = 0) and those who were lost to follow up (LTFU = 1), at the following levels: *p<0.05, **p<0.01, ***p<0.001.

**(b) Pulmonary and Peripheral Airway Function**

| | **Retained** | **LTFU** |
|---|---|---|
| **Pulmonary Function** | | |
| $FEV_1$ %pred | 103 ± 13 | 110 ± 12 |
| $FEV_1$/FVC (%) | 79 ± 6 (68-90) | 79 ± 6 (67-90) |
| $FEF_{25-75}$ (%) | 84 ± 29 | 97 ± 26 |
| TLC %pred | 100 ± 9 | 98 ± 10 |
| RV/TLC | 31 ± 5.6 | 27 ± 6.2 |
| DLCO %pred | 79 ± 13 | 85 ± 15 |
| Lung Function Score | 2.9 ± 0.88 | 2.6 ± 0.78 |
| **SGRQ Total Score** | 12 ± 11 | 14 ± 10 |
| **Peripheral Airway Function** | | |
| $S_{acin}$ ($L^{-1}$) | 0.08 ± 0.04 | 0.114 ± 0.05* |
| $S_{cond}$ ($L^{-1}$) | 0.03 ± 0.01 | 0.033 ± 0.02 |
| LCI | 8.65 ± 1.15 | 8.52 ± 1.07 |
| Rrs (cmH20) | 3.70 ± 1.47 | 3.47 ± 1.13 |
| Xrs (cmH20) | -0.67 ± 0.56 | -0.65 ± 0.46 |

Footnote: Data displayed as Mean±SD or N (%) unless otherwise stated. Asterisks indicate a statistically significant difference, in either the mean or proportion (%) of pulmonary and peripheral function parameters, between those who completed their study visits (LTFU = 0) and those who were lost to follow up (LTFU = 1), at the following levels: *p<0.05, **p<0.01, ***p<0.001.

**3. Changes in lung function outcomes**

**3.1 Changes in lung function outcomes across the entire cohort for the 3 year follow up period**

**Figure E1**. Change in $S_{acin}$, LCI and $FEV_1$ across the three year follow up period post HSCT, for the entire cohort.

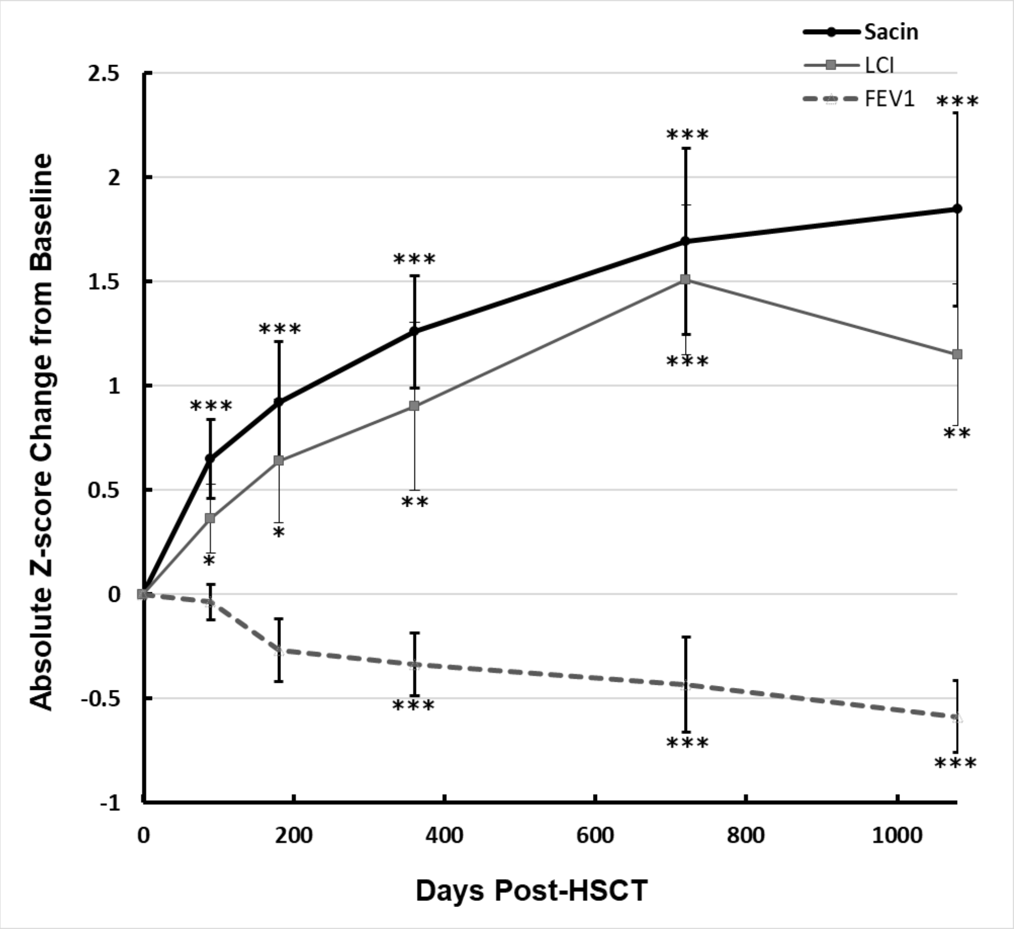

**Table E5**. Estimated rates of Z-score change per month in MBNW, oscillometry and spirometry parameters across the 3 year period post HSCT, across the entire cohort.

| **Days Post-HSCT** | **MBW Parameter** | ***All Subjects*** | | | ***Oscillometry Parameter*** | ***All Subjects*** | | | ***Spirometry Parameter*** | ***All Subjects*** | | |
|---|---|---|---|---|---|---|---|---|---|---|---|---|
| | | **estimate*** | **S.E.** | **p value** | | **estimate*** | **S.E.** | **p value** | | **estimate*** | **S.E.** | **p value** |
| **90** | $S_{acin}$ Z-score | 0.224 | 0.05 | **<0.001** | Rrs Z-score | -0.131 | 0.12 | 0.27 | $FEV_1$ Z-score | -0.023 | 0.02 | 0.33 |
| | $S_{cond}$ Z-score | 0.020 | 0.05 | 0.70 | Xrs Z-score | 0.005 | 0.07 | 0.94 | $FEV_1$/FVC Z-score | 0.026 | 0.03 | 0.40 |
| | LCI Z-score | 0.126 | 0.05 | **0.03** | | | | | $FEF_{25-75}$ Z-score | -0.005 | 0.03 | 0.84 |
| **180** | $S_{acin}$ Z-score | 0.181 | 0.03 | **<0.001** | Rrs Z-score | -0.097 | 0.07 | 0.15 | $FEV_1$ Z-score | -0.034 | 0.02 | 0.06 |
| | $S_{cond}$ Z-score | -0.021 | 0.03 | 0.50 | Xrs Z-score | -0.054 | 0.04 | 0.21 | $FEV_1$/FVC Z-score | 0.026 | 0.02 | 0.13 |
| | LCI Z-score | 0.089 | 0.04 | **0.03** | | | | | $FEF_{25-75}$ Z-score | -0.004 | 0.02 | 0.81 |
| **270** | $S_{acin}$ Z-score | 0.124 | 0.02 | **<0.001** | Rrs Z-score | -0.083 | 0.05 | 0.07 | $FEV_1$ Z-score | -0.027 | 0.01 | **0.02** |
| | $S_{cond}$ Z-score | -0.003 | 0.02 | 0.89 | Xrs Z-score | -0.040 | 0.03 | 0.23 | $FEV_1$/FVC Z-score | 0.022 | 0.01 | 0.09 |
| | LCI Z-score | 0.020 | 0.03 | 0.43 | | | | | $FEF_{25-75}$ Z-score | 0.002 | 0.01 | 0.87 |
| **360** | $S_{acin}$ Z-score | 0.113 | 0.01 | **<0.001** | Rrs Z-score | -0.040 | 0.03 | 0.18 | $FEV_1$ Z-score | -0.033 | 0.01 | **<0.001** |
| | $S_{cond}$ Z-score | -2.5E-06 | 0.01 | 1 | Xrs Z-score | -0.035 | 0.03 | 0.17 | $FEV_1$/FVC Z-score | -0.003 | 0.01 | 0.74 |
| | LCI Z-score | 0.059 | 0.02 | **0.006** | | | | | $FEF_{25-75}$ Z-score | -0.013 | 0.01 | 0.15 |
| **450** | $S_{acin}$ Z-score | 0.093 | 0.01 | **<0.001** | Rrs Z-score | 0.001 | 0.03 | 0.97 | $FEV_1$ Z-score | -0.030 | 0.01 | **<0.001** |
| | $S_{cond}$ Z-score | 0.016 | 0.01 | 0.26 | Xrs Z-score | -0.039 | 0.02 | 0.10 | $FEV_1$/FVC Z-score | -0.006 | 0.01 | 0.44 |
| | LCI Z-score | 0.040 | 0.02 | **0.04** | | | | | $FEF_{25-75}$ Z-score | -0.017 | 0.01 | **0.04** |
| **540** | $S_{acin}$ Z-score | 0.081 | 0.01 | **<0.001** | Rrs Z-score | -0.002 | 0.02 | 0.94 | $FEV_1$ Z-score | -0.025 | 0.01 | **<0.001** |
| | $S_{cond}$ Z-score | 0.021 | 0.01 | 0.07 | Xrs Z-score | -0.018 | 0.02 | 0.35 | $FEV_1$/FVC Z-score | -0.012 | 0.01 | 0.07 |
| | LCI Z-score | 0.036 | 0.02 | **0.03** | | | | | $FEF_{25-75}$ Z-score | -0.018 | 0.01 | **0.008** |
| **630** | $S_{acin}$ Z-score | 0.085 | 0.01 | **<0.001** | Rrs Z-score | -0.002 | 0.02 | 0.92 | $FEV_1$ Z-score | -0.027 | 0.01 | **<0.001** |
| | $S_{cond}$ Z-score | 0.032 | 0.01 | **0.002** | Xrs Z-score | -0.032 | 0.02 | 0.12 | $FEV_1$/FVC Z-score | -0.017 | 0.01 | **0.007** |
| | LCI Z-score | 0.058 | 0.02 | **0.001** | | | | | $FEF_{25-75}$ Z-score | -0.021 | 0.01 | **0.001** |
| **720** | $S_{acin}$ Z-score | 0.074 | 0.01 | **<0.001** | Rrs Z-score | 0.018 | 0.02 | 0.32 | $FEV_1$ Z-score | -0.022 | 0.01 | **<0.001** |

| | | | | | | | | | | | | |
|---|---|---|---|---|---|---|---|---|---|---|---|---|
| | $S_{cond}$ Z-score | 0.042 | 0.01 | **<0.001** | Xrs Z-score | -0.039 | 0.02 | **0.03** | $FEV_1$/FVC Z-score | -0.020 | 0.01 | **0.001** |
| | LCI Z-score | 0.063 | 0.01 | **<0.001** | | | | | $FEF_{25-75}$ Z-score | -0.020 | 0.01 | **<0.001** |
| **810** B | $S_{acin}$ Z-score | 0.062 | 0.01 | **<0.001** | Rrs Z-score | 0.017 | 0.02 | 0.25 | $FEV_1$ Z-score | -0.017 | 0.005 | **<0.001** |
| | $S_{cond}$ Z-score | 0.034 | 0.01 | **<0.001** | Xrs Z-score | -0.031 | 0.01 | **0.04** | $FEV_1$/FVC Z-score | -0.018 | 0.005 | **<0.001** |
| | LCI Z-score | 0.051 | 0.01 | **<0.001** | | | | | $FEF_{25-75}$ Z-score | -0.017 | 0.005 | **<0.001** |
| **900** | $S_{acin}$ Z-score | 0.053 | 0.01 | **<0.001** | Rrs Z-score | 0.024 | 0.01 | 0.08 | $FEV_1$ Z-score | -0.015 | 0.004 | **<0.001** |
| | $S_{cond}$ Z-score | 0.030 | 0.01 | **<0.001** | Xrs Z-score | -0.029 | 0.01 | **0.03** | $FEV_1$/FVC Z-score | -0.016 | 0.004 | **<0.001** |
| | LCI Z-score | 0.043 | 0.01 | **<0.001** | | | | | $FEF_{25-75}$ Z-score | -0.016 | 0.004 | **<0.001** |
| **990** | $S_{acin}$ Z-score | 0.048 | 0.01 | **<0.001** | Rrs Z-score | 0.018 | 0.01 | 0.15 | $FEV_1$ Z-score | -0.013 | 0.004 | **0.001** |
| | $S_{cond}$ Z-score | 0.027 | 0.01 | **<0.001** | Xrs Z-score | -0.021 | 0.01 | 0.08 | $FEV_1$/FVC Z-score | -0.015 | 0.004 | **<0.001** |
| | LCI Z-score | 0.037 | 0.01 | **0.001** | | | | | $FEF_{25-75}$ Z-score | -0.015 | 0.004 | **<0.001** |
| **1080** | $S_{acin}$ Z-score | 0.045 | 0.01 | **<0.001** | Rrs Z-score | 0.021 | 0.01 | 0.06 | $FEV_1$ Z-score | -0.012 | 0.003 | **<0.001** |
| | $S_{cond}$ Z-score | 0.024 | 0.01 | **<0.001** | Xrs Z-score | -0.024 | 0.01 | **0.03** | $FEV_1$/FVC Z-score | -0.013 | 0.003 | **<0.001** |
| | LCI Z-score | 0.032 | 0.01 | **0.001** | | | | | $FEF_{25-75}$ Z-score | -0.014 | 0.003 | **<0.001** |

Footnote: *Mixed model estimates represent the rate of change in Z-score per month; S.E., standard error.

**3.2 Change in lung function parameters across the groups stratified by BOS 0-p status**

The mixed model estimates for the absolute Z-score change in $S_{acin}$, $FEV_1$ and $FEF_{25-75}$ from baseline, with days post-HSCT within the first year, in the BOS-0p (a) and No BOS-0p (b) groups are provided in Table E6. Further details of change in all measured parameters, over the entire follow-up period of 36 months, are provided in Table E7.

**Table E6**. Estimated rates of Z-score change per month in $S_{acin}$, $FEV_1$ and $FEF_{25-75}$ within the first year post-HSCT, in groups stratified by BOS-0p status.

| Days Post-HSCT | Parameter | *BOS-0p* | | | *NO BOS-0p* | | |
|---|---|---|---|---|---|---|---|
| | | **estimate*** | **S.E.** | **p value** | **estimate*** | **S.E.** | **p value** |
| **90**<br>BOS-0p n=11<br>No BOS-0p n=27 | $S_{acin}$ Z-score | 0.28 | 0.12 | 0.03 | 0.15 | 0.03 | <0.001 |
| | $FEV_1$ Z-score | -0.05 | 0.04 | 0.29 | 0.002 | 0.05 | 0.95 |
| | $FEF_{25-75}$ Z-score | -0.06 | 0.07 | 0.41 | 0.03 | 0.02 | 0.22 |
| **180**<br>BOS-0p n=14<br>No BOS-0p n=16 | $S_{acin}$ Z-score | 0.21 | 0.06 | <0.001 | 0.14 | 0.04 | <0.001 |
| | $FEV_1$ Z-score | -0.09 | 0.03 | 0.007 | 0.01 | 0.02 | 0.49 |
| | $FEF_{25-75}$ Z-score | -0.05 | 0.03 | 0.12 | 0.04 | 0.01 | 0.01 |
| **360**<br>BOS-0p n=14<br>No BOS-0p n=19 | $S_{acin}$ Z-score | 0.16 | 0.02 | <0.001 | 0.08 | 0.02 | <0.001 |
| | $FEV_1$ Z-score | -0.10 | 0.01 | <0.001 | 0.02 | 0.01 | 0.08 |
| | $FEF_{25-75}$ Z-score | -0.08 | 0.01 | <0.001 | 0.04 | 0.01 | <0.001 |

Footnote: *Mixed model estimates represent the rate of change in Z-score per month; S.E., standard error

**Table E7**. Estimated rates of Z-score change per month in MBW and oscillometry parameters, across the 3 year period post HSCT, in groups stratified by BOS-0p status.

(a)

| Days Post HSCT | MBW Parameter | BOS-0p | | | No BOS-0p | | | Oscillometry Parameter | BOS-0p | | | No BOS-0p | | |
|---|---|---|---|---|---|---|---|---|---|---|---|---|---|---|
| | | **estimate*** | **S.E.** | **p value** | **estimate*** | **S.E.** | **p value** | | **estimate*** | **S.E.** | **p value** | **estimate*** | **S.E.** | **p value** |
| **90** BOS-0p n=12 No BOS-0p n=27 | $S_{acin}$ Z-score | 0.335 | 0.12 | **0.01** | 0.153 | 0.03 | **<0.001** | Rrs Z-score | -0.103 | 0.21 | 0.64 | -0.162 | 0.14 | 0.27 |
| | $S_{cond}$ Z-score | 0.138 | 0.08 | 0.09 | -0.035 | 0.06 | 0.58 | Xrs Z-score | 0.029 | 0.13 | 0.83 | -0.016 | 0.08 | 0.84 |
| | LCI Z-score | 0.104 | 0.07 | 0.17 | 0.127 | 0.07 | 0.10 | | | | | | | |
| **180** BOS-0p n= 15 No BOS-0p n= 16 | $S_{acin}$ Z-score | 0.224 | 0.06 | **<0.001** | 0.140 | 0.04 | **<0.001** | Rrs Z-score | 0.012 | 0.10 | 0.91 | -0.190 | 0.09 | 0.04 |
| | $S_{cond}$ Z-score | -0.012 | 0.04 | 0.79 | -0.021 | 0.04 | 0.62 | Xrs Z-score | -0.133 | 0.07 | 0.07 | 0.002 | 0.05 | 0.97 |
| | LCI Z-score | 0.128 | 0.06 | **0.03** | 0.055 | 0.05 | 0.31 | | | | | | | |
| **270** BOS-0p n= 13 No BOS-0p n= 19 | $S_{acin}$ Z-score | 0.175 | 0.04 | **<0.001** | 0.086 | 0.02 | **<0.001** | Rrs Z-score | 0.022 | 0.08 | 0.77 | -0.152 | 0.06 | 0.008 |
| | $S_{cond}$ Z-score | -0.016 | 0.03 | 0.57 | 0.009 | 0.03 | 0.72 | Xrs Z-score | -0.116 | 0.06 | 0.07 | 0.003 | 0.04 | 0.94 |
| | LCI Z-score | 0.052 | 0.04 | 0.16 | -0.005 | 0.04 | 0.88 | | | | | | | |
| **360** BOS-0p n= 14 No BOS-0p n= 19 | $S_{acin}$ Z-score | 0.157 | 0.02 | **<0.001** | 0.078 | 0.02 | **<0.001** | Rrs Z-score | 0.022 | 0.05 | 0.64 | -0.093 | 0.04 | 0.02 |
| | $S_{cond}$ Z-score | 0.020 | 0.02 | 0.36 | -0.015 | 0.02 | 0.42 | Xrs Z-score | -0.107 | 0.05 | **0.03** | 0.021 | 0.03 | 0.42 |
| | LCI Z-score | 0.147 | 0.03 | **<0.001** | -0.011 | 0.03 | 0.66 | | | | | | | |
| **450** BOS-0p n= 13 No BOS-0p n= 9 | $S_{acin}$ Z-score | 0.126 | 0.02 | **<0.001** | 0.060 | 0.01 | **<0.001** | Rrs Z-score | 0.015 | 0.04 | 0.68 | -0.024 | 0.04 | 0.57 |
| | $S_{cond}$ Z-score | 0.045 | 0.02 | 0.06 | -0.013 | 0.02 | 0.42 | Xrs Z-score | -0.091 | 0.04 | **0.02** | 0.011 | 0.03 | 0.69 |
| | LCI Z-score | 0.112 | 0.03 | **0.001** | -0.028 | 0.02 | 0.19 | | | | | | | |
| **540** BOS-0p n= 13 No BOS-0p n= 13 | $S_{acin}$ Z-score | 0.121 | 0.02 | **<0.001** | 0.045 | 0.01 | **<0.001** | Rrs Z-score | -0.008 | 0.03 | 0.77 | -0.003 | 0.03 | 0.93 |
| | $S_{cond}$ Z-score | 0.053 | 0.02 | **0.007** | -0.009 | 0.01 | 0.46 | Xrs Z-score | -0.039 | 0.03 | 0.19 | 0.002 | 0.02 | 0.94 |
| | LCI Z-score | 0.109 | 0.03 | **<0.001** | -0.032 | 0.02 | 0.05 | | | | | | | |
| **630** BOS-0p n= 6 No BOS-0p n= 8 | $S_{acin}$ Z-score | 0.140 | 0.02 | **<0.001** | 0.035 | 0.01 | **0.001** | Rrs Z-score | 0.007 | 0.03 | 0.79 | -0.017 | 0.03 | 0.58 |
| | $S_{cond}$ Z-score | 0.072 | 0.02 | **<0.001** | -0.003 | 0.01 | 0.74 | Xrs Z-score | -0.078 | 0.04 | **0.04** | 0.007 | 0.02 | 0.77 |

| | | | | | | | | | | | | | | |
|---|---|---|---|---|---|---|---|---|---|---|---|---|---|---|
| | LCI Z-score | 0.148 | 0.03 | **<0.001** | -0.026 | 0.01 | 0.08 | | | | | | | |
| **720**<br>BOS-0p n= 7<br>No BOS-0p n= 11 | $S_{acin}$ Z-score | 0.121 | 0.02 | **<0.001** | 0.035 | 0.01 | **<0.001** | Rrs Z-score | 0.016 | 0.02 | 0.49 | 0.014 | 0.03 | 0.59 |
| | $S_{cond}$ Z-score | 0.095 | 0.02 | **<0.001** | -0.003 | 0.01 | 0.72 | Xrs Z-score | -0.067 | 0.03 | **0.04** | -0.018 | 0.02 | 0.35 |
| | LCI Z-score | 0.126 | 0.02 | **<0.001** | 0.008 | 0.02 | 0.62 | | | | | | | |
| **810**<br>BOS-0p n= 6<br>No BOS-0p n= 13 | $S_{acin}$ Z-score | 0.103 | 0.02 | **<0.001** | 0.033 | 0.01 | **<0.001** | Rrs Z-score | 0.016 | 0.02 | 0.45 | 0.013 | 0.02 | 0.53 |
| | $S_{cond}$ Z-score | 0.072 | 0.02 | **<0.001** | 0.008 | 0.01 | 0.28 | Xrs Z-score | -0.047 | 0.03 | 0.10 | -0.020 | 0.02 | 0.20 |
| | LCI Z-score | 0.117 | 0.02 | **<0.001** | 0.002 | 0.01 | 0.87 | | | | | | | |
| **900**<br>BOS-0p n= 9<br>No BOS-0p n= 5 | $S_{acin}$ Z-score | 0.081 | 0.01 | **<0.001** | 0.029 | 0.01 | **<0.001** | Rrs Z-score | 0.020 | 0.02 | 0.26 | 0.022 | 0.02 | 0.27 |
| | $S_{cond}$ Z-score | 0.060 | 0.01 | **<0.001** | 0.005 | 0.01 | 0.46 | Xrs Z-score | -0.037 | 0.02 | 0.13 | -0.022 | 0.01 | 0.13 |
| | LCI Z-score | 0.094 | 0.02 | **<0.001** | -0.002 | 0.01 | 0.88 | | | | | | | |
| **990**<br>BOS-0p n= 7<br>No BOS-0p n= 7 | $S_{acin}$ Z-score | 0.069 | 0.01 | **<0.001** | 0.029 | 0.01 | **<0.001** | Rrs Z-score | 0.014 | 0.02 | 0.38 | 0.016 | 0.02 | 0.37 |
| | $S_{cond}$ Z-score | 0.052 | 0.01 | **<0.001** | 0.006 | 0.01 | 0.38 | Xrs Z-score | -0.030 | 0.02 | 0.17 | -0.014 | 0.01 | 0.28 |
| | LCI Z-score | 0.081 | 0.02 | **<0.001** | -0.002 | 0.01 | 0.83 | | | | | | | |
| **1080**<br>BOS-0p n= 8<br>No BOS-0p n= 7 | $S_{acin}$ Z-score | 0.061 | 0.01 | **<0.001** | 0.030 | 0.01 | **<0.001** | Rrs Z-score | 0.016 | 0.01 | 0.27 | 0.023 | 0.02 | 0.18 |
| | $S_{cond}$ Z-score | 0.050 | 0.01 | **<0.001** | -0.0001 | 0.01 | 0.99 | Xrs Z-score | -0.035 | 0.02 | 0.07 | -0.014 | 0.01 | 0.27 |
| | LCI Z-score | 0.067 | 0.02 | **<0.001** | -0.001 | 0.01 | 0.90 | | | | | | | |

Footnote: *Mixed model estimates represent the rate of change in Z-score per month; S.E., standard error

##### 3.3 Change in lung function parameters across the first year stratified by cGVHD status

The mixed model estimates for the absolute Z-score change in $S_{acin}$, $FEV_1$ and $FEF_{25-75}$ from baseline, with days post-HSCT within the first year, in the (a) cGVHD and (b) No cGVHD groups are provided in Table E8, and in Table E9.

**Table E8**. Estimated rates of Z-score change per month in $S_{acin}$, LCI and $FEV_1$ within the first year post-HSCT, in groups stratified by cGVHD status.

| **Days Post HSCT** | **Parameter** | ***cGVHD Group*** | | | ***No-cGVHD Group*** | | |
|---|---|---|---|---|---|---|---|
| | | **estimate*** | **S.E.** | **p value** | **estimate*** | **S.E.** | **p value** |
| **90**<br>cGVHD n= 27<br>No cGVHD n= 11 | $S_{acin}$ Z-score | 0.25 | 0.06 | 0.001 | 0.09 | 0.05 | 0.09 |
| | LCI Z-score | 0.21 | 0.07 | 0.01 | -0.01 | 0.08 | 0.90 |
| | $FEV_1$ Z-score | -0.01 | 0.03 | 0.64 | -0.02 | 0.04 | 0.67 |
| **180**<br>cGVHD n= 21<br>No cGVHD n= 9 | $S_{acin}$ Z-score | 0.22 | 0.04 | <0.001 | 0.06 | 0.03 | 0.10 |
| | LCI Z-score | 0.15 | 0.05 | 0.01 | -0.03 | 0.05 | 0.51 |
| | $FEV_1$ Z-score | -0.03 | 0.02 | 0.13 | -0.03 | 0.04 | 0.43 |
| **360**<br>cGVHD n= 23<br>No cGVHD n= 10 | $S_{acin}$ Z-score | 0.14 | 0.02 | <0.001 | 0.03 | 0.02 | 0.09 |
| | LCI Z-score | 0.11 | 0.03 | <0.001 | -0.06 | 0.02 | 0.02 |
| | $FEV_1$ Z-score | -0.04 | 0.01 | <0.001 | -0.01 | 0.02 | 0.73 |

Footnote: *Mixed model estimates represent the rate of change in Z-score per month; S.E., standard error

**Table E9**. Estimated rates of change in (a) MBW, (b) oscillometry and (c) spirometry parameters, across the 3 year period post HSCT, in groups stratified by cGVHD status.

**A.**

| Days Post-HSCT | Parameter | cGVHD | | | No cGVHD | | |
|---|---|---|---|---|---|---|---|
| | | estimate* | S.E. | p value | estimate* | S.E. | p value |
| **90** cGVHD n= 27 No cGVHD n= 11 | $S_{acin}$ Z-score | 0.249 | 0.06 | **0.001** | 0.093 | 0.05 | 0.09 |
| | $S_{cond}$ Z-score | 0.048 | 0.07 | 0.46 | -0.064 | 0.08 | 0.43 |
| | LCI Z-score | 0.209 | 0.07 | **0.01** | -0.011 | 0.08 | 0.90 |
| **180** cGVHD n= 21 No cGVHD n= 9 | $S_{acin}$ Z-score | 0.216 | 0.04 | **<0.001** | 0.058 | 0.03 | 0.10 |
| | $S_{cond}$ Z-score | -0.001 | 0.04 | 0.99 | -0.071 | 0.04 | 0.09 |
| | LCI Z-score | 0.147 | 0.05 | **0.01** | -0.034 | 0.05 | 0.51 |
| **270** cGVHD n= 22 No cGVHD n= 10 | $S_{acin}$ Z-score | 0.146 | 0.03 | **<0.001** | 0.056 | 0.02 | 0.28 |
| | $S_{cond}$ Z-score | 0.008 | 0.02 | 0.74 | -0.032 | 0.03 | 0.31 |
| | LCI Z-score | 0.058 | 0.03 | 0.09 | -0.066 | 0.03 | 0.048 |
| **360** cGVHD n= 23 No cGVHD n= 10 | $S_{acin}$ Z-score | 0.142 | 0.02 | **<0.001** | 0.030 | 0.02 | 0.09 |
| | $S_{cond}$ Z-score | 0.012 | 0.02 | 0.70 | -0.028 | 0.02 | 0.22 |
| | LCI Z-score | 0.111 | 0.03 | **<0.001** | -0.056 | 0.02 | 0.02 |
| **450** cGVHD n= 16 No cGVHD n= 6 | $S_{acin}$ Z-score | 0.121 | 0.01 | **<0.001** | 0.020 | 0.02 | 0.26 |
| | $S_{cond}$ Z-score | 0.031 | 0.02 | 0.08 | -0.021 | 0.02 | 0.26 |
| | LCI Z-score | 0.083 | 0.03 | **0.002** | -0.053 | 0.02 | 0.008 |
| **540** cGVHD n= 16 No cGVHD n= 10 | $S_{acin}$ Z-score | 0.106 | 0.01 | **<0.001** | 0.029 | 0.01 | **0.03** |
| | $S_{cond}$ Z-score | 0.036 | 0.01 | **0.02** | -0.010 | 0.02 | 0.51 |
| | LCI Z-score | 0.062 | 0.02 | **0.008** | -0.012 | 0.02 | 0.50 |
| **630** cGVHD n= 9 No cGVHD n= 5 | $S_{acin}$ Z-score | 0.110 | 0.02 | **<0.001** | 0.030 | 0.01 | **0.02** |
| | $S_{cond}$ Z-score | 0.046 | 0.01 | **0.001** | 0.003 | 0.01 | 0.82 |
| | LCI Z-score | 0.080 | 0.02 | **<0.001** | 0.017 | 0.02 | 0.46 |
| **720** cGVHD n= 12 No cGVHD n= 6 | $S_{acin}$ Z-score | 0.099 | 0.01 | **<0.001** | 0.022 | 0.01 | 0.05 |
| | $S_{cond}$ Z-score | 0.046 | 0.01 | **<0.001** | 0.035 | 0.02 | 0.07 |
| | LCI Z-score | 0.090 | 0.02 | **<0.001** | 0.011 | 0.02 | 0.57 |
| **810** cGVHD n= 13 No cGVHD n= 6 | $S_{acin}$ Z-score | 0.084 | 0.01 | **<0.001** | 0.018 | 0.01 | 0.05 |
| | $S_{cond}$ Z-score | 0.037 | 0.01 | **<0.001** | 0.030 | 0.02 | 0.06 |
| | LCI Z-score | 0.078 | 0.02 | **<0.001** | -0.003 | 0.02 | 0.87 |
| **900** cGVHD n= 12 No cGVHD n= 2 | $S_{acin}$ Z-score | 0.069 | 0.01 | **<0.001** | 0.016 | 0.01 | 0.07 |
| | $S_{cond}$ Z-score | 0.031 | 0.01 | **0.001** | 0.029 | 0.01 | 0.05 |
| | LCI Z-score | 0.065 | 0.02 | **<0.001** | -0.007 | 0.02 | 0.64 |
| **990** cGVHD n= 9 No cGVHD n= 5 | $S_{acin}$ Z-score | 0.064 | 0.01 | **<0.001** | 0.011 | 0.01 | 0.16 |
| | $S_{cond}$ Z-score | 0.031 | 0.01 | **<0.001** | 0.018 | 0.01 | 0.18 |
| | LCI Z-score | 0.061 | 0.01 | **<0.001** | -0.016 | 0.01 | 0.26 |
| **1080** cGVHD n= 10 No cGVHD n= 5 | $S_{acin}$ Z-score | 0.058 | 0.01 | **<0.001** | 0.014 | 0.01 | 0.05 |
| | $S_{cond}$ Z-score | 0.032 | 0.01 | **<0.001** | 0.0051 | 0.01 | 0.68 |
| | LCI Z-score | 0.054 | 0.01 | **<0.001** | -0.016 | 0.01 | 0.20 |

Footnote: *Mixed model estimates represent the rate of change in Z-score per month; S.E., standard error

**B.**

| Days Post-HSCT | Parameter | cGVHD | | | No cGVHD | | |
|---|---|---|---|---|---|---|---|
| | | estimate* | S.E. | p value | estimate* | S.E. | p value |
| **90** cGVHD n= 27 No cGVHD n= 11 | Rrs Z-score | -0.056 | 0.11 | 0.63 | -0.312 | 0.29 | 0.30 |
| | Xrs Z-score | 0.027 | 0.08 | 0.74 | -0.008 | 0.13 | 0.95 |
| **180** cGVHD n= 21 No cGVHD n= 9 | Rrs Z-score | -0.049 | 0.06 | 0.43 | -0.227 | 0.17 | 0.20 |
| | Xrs Z-score | -0.035 | 0.05 | 0.46 | -0.069 | 0.09 | 0.45 |
| **270** cGVHD n= 22 No cGVHD n= 10 | Rrs Z-score | -0.053 | 0.04 | 0.22 | -0.170 | 0.12 | 0.160 |
| | Xrs Z-score | -0.055 | 0.04 | 0.16 | 0.012 | 0.06 | 0.85 |
| **360** cGVHD n= 23 No cGVHD n= 10 | Rrs Z-score | -0.015 | 0.03 | 0.61 | -0.127 | 0.08 | 0.11 |
| | Xrs Z-score | -0.064 | 0.03 | 0.05 | 0.051 | 0.04 | 0.22 |
| **450** cGVHD n=16 No cGVHD n= 6 | Rrs Z-score | 0.004 | 0.03 | 0.87 | -0.025 | 0.08 | 0.75 |
| | Xrs Z-score | -0.058 | 0.03 | **0.048** | 0.014 | 0.04 | 0.70 |
| **540** cGVHD n= 16 No cGVHD n= 10 | Rrs Z-score | 0.004 | 0.02 | 0.85 | -0.019 | 0.06 | 0.74 |
| | Xrs Z-score | -0.020 | 0.02 | 0.42 | -0.012 | 0.03 | 0.68 |
| **630** cGVHD n= 9 No cGVHD n= 5 | Rrs Z-score | 0.016 | 0.02 | 0.42 | -0.042 | 0.05 | 0.38 |
| | Xrs Z-score | -0.048 | 0.03 | 0.09 | -0.004 | 0.03 | 0.86 |
| **720** cGVHD n= 12 No cGVHD n= 6 | Rrs Z-score | 0.016 | 0.02 | 0.35 | 0.020 | 0.04 | 0.65 |
| | Xrs Z-score | -0.040 | 0.02 | 0.09 | -0.040 | 0.02 | 0.09 |
| **810** cGVHD n= 13 No cGVHD n= 6 | Rrs Z-score | 0.014 | 0.01 | 0.33 | 0.021 | 0.04 | 0.56 |
| | Xrs Z-score | -0.031 | 0.02 | 0.12 | -0.032 | 0.02 | 0.10 |
| **900** cGVHD n= 12 No cGVHD n= 2 | Rrs Z-score | 0.025 | 0.01 | 0.05 | 0.018 | 0.03 | 0.60 |
| | Xrs Z-score | -0.031 | 0.02 | 0.07 | -0.025 | 0.02 | 0.16 |
| **990** cGVHD n= 9 No cGVHD n= 5 | Rrs Z-score | 0.021 | 0.01 | 0.08 | 0.008 | 0.03 | 0.79 |
| | Xrs Z-score | -0.026 | 0.02 | 0.10 | -0.013 | 0.02 | 0.42 |
| **1080** cGVHD n= 10 No cGVHD n= 5 | Rrs Z-score | 0.023 | 0.01 | **0.03** | 0.016 | 0.03 | 0.58 |
| | Xrs Z-score | -0.029 | 0.01 | 0.05 | -0.015 | 0.02 | 0.32 |

Footnote: *Mixed model estimates represent the rate of change in Z-score per month; S.E., standard error

**C.**

| Days Post-HSCT | Parameter | cGVHD | | | No cGVHD | | |
|---|---|---|---|---|---|---|---|
| | | estimate* | S.E. | p value | estimate* | S.E. | p value |
| **90** cGVHD n= 27 No cGVHD n= 11 | $FEV_1$ Z-score | -0.014 | 0.03 | 0.64 | -0.016 | 0.04 | 0.67 |
| | $FEV_1$/FVC Z-score | 0.020 | 0.04 | 0.59 | 0.035 | 0.06 | 0.59 |
| | $FEF_{25-75}$ Z-score | -0.003 | 0.03 | 0.93 | 0.009 | 0.04 | 0.83 |
| **180** cGVHD n= 21 No cGVHD n= 9 | $FEV_1$ Z-score | -0.032 | 0.02 | 0.13 | -0.028 | 0.04 | 0.43 |
| | $FEV_1$/FVC Z-score | 0.029 | 0.02 | 0.16 | 0.019 | 0.04 | 0.60 |
| | $FEF_{25-75}$ Z-score | 0.009 | 0.02 | 0.60 | -0.027 | 0.03 | 0.41 |
| **270** cGVHD n= 22 No cGVHD n= 10 | $FEV_1$ Z-score | -0.025 | 0.01 | 0.07 | -0.026 | 0.02 | 0.26 |
| | $FEV_1$/FVC Z-score | 0.031 | 0.01 | 0.04 | 0.006 | 0.02 | 0.80 |
| | $FEF_{25-75}$ Z-score | 0.011 | 0.01 | 0.36 | -0.014 | 0.02 | 0.50 |
| **360** cGVHD n= 23 No cGVHD n= 10 | $FEV_1$ Z-score | -0.044 | 0.01 | **<0.001** | -0.006 | 0.02 | 0.73 |
| | $FEV_1$/FVC Z-score | -0.004 | 0.01 | 0.71 | 0.001 | 0.02 | 0.95 |
| | $FEF_{25-75}$ Z-score | -0.014 | 0.01 | 0.21 | -0.010 | 0.01 | 0.47 |
| **450** cGVHD n= 16 No cGVHD n= 6 | $FEV_1$ Z-score | -0.040 | 0.01 | **<0.001** | -0.005 | 0.01 | 0.71 |
| | $FEV_1$/FVC Z-score | -0.009 | 0.01 | 0.37 | 0.001 | 0.01 | 0.94 |
| | $FEF_{25-75}$ Z-score | -0.022 | 0.01 | **0.04** | -0.008 | 0.01 | 0.46 |
| **540** cGVHD n= 16 No cGVHD n= 10 | $FEV_1$ Z-score | -0.034 | 0.01 | **<0.001** | -0.007 | 0.01 | 0.51 |
| | $FEV_1$/FVC Z-score | -0.012 | 0.01 | 0.17 | -0.012 | 0.01 | 0.21 |
| | $FEF_{25-75}$ Z-score | -0.021 | 0.01 | **0.02** | -0.012 | 0.01 | 0.14 |
| **630** cGVHD n= 9 No cGVHD n= 5 | $FEV_1$ Z-score | -0.039 | 0.01 | **<0.001** | -0.002 | 0.01 | 0.79 |
| | $FEV_1$/FVC Z-score | -0.021 | 0.01 | **0.01** | -0.008 | 0.01 | 0.34 |
| | $FEF_{25-75}$ Z-score | -0.030 | 0.01 | **<0.001** | -0.006 | 0.01 | 0.42 |
| **720** cGVHD n= 12 No cGVHD n= 6 | $FEV_1$ Z-score | -0.033 | 0.01 | **<0.001** | -0.001 | 0.01 | 0.89 |
| | $FEV_1$/FVC Z-score | -0.022 | 0.01 | **0.002** | -0.015 | 0.01 | 0.09 |
| | $FEF_{25-75}$ Z-score | -0.027 | 0.01 | **<0.001** | -0.005 | 0.01 | 0.41 |
| **810** cGVHD n= 13 No cGVHD n= 6 | $FEV_1$ Z-score | -0.027 | 0.01 | **<0.001** | 0.002 | 0.01 | 0.72 |
| | $FEV_1$/FVC Z-score | -0.021 | 0.01 | **0.001** | -0.011 | 0.01 | 0.15 |
| | $FEF_{25-75}$ Z-score | -0.024 | 0.01 | **<0.001** | -0.003 | 0.01 | 0.56 |
| **900** cGVHD n= 12 No cGVHD n= 2 | $FEV_1$ Z-score | -0.024 | 0.01 | **<0.001** | 0.004 | 0.01 | 0.47 |
| | $FEV_1$/FVC Z-score | -0.020 | 0.01 | **<0.001** | -0.008 | 0.01 | 0.23 |
| | $FEF_{25-75}$ Z-score | -0.022 | 0.01 | **<0.001** | -0.004 | 0.01 | 0.39 |
| **990** cGVHD n= 9 No cGVHD n= 5 | $FEV_1$ Z-score | -0.022 | 0.01 | **<0.001** | 0.006 | 0.01 | 0.20 |
| | $FEV_1$/FVC Z-score | -0.019 | 0.01 | **<0.001** | -0.006 | 0.01 | 0.36 |
| | $FEF_{25-75}$ Z-score | -0.022 | 0.01 | **<0.001** | -0.002 | 0.01 | 0.58 |
| **1080** cGVHD n= 10 No cGVHD n= 5 | $FEV_1$ Z-score | -0.020 | 0.01 | **<0.001** | 0.005 | 0.01 | 0.26 |
| | $FEV_1$/FVC Z-score | -0.018 | 0.01 | **<0.001** | -0.004 | 0.01 | 0.48 |
| | $FEF_{25-75}$ Z-score | -0.021 | 0.01 | **<0.001** | -0.001 | 0.01 | 0.82 |

Footnote: *Mixed model estimates represent the rate of change in Z-score per month; S.E., standard error

**Table E10.** Absolute change from baseline in $S_{acin}$ between groups stratified by cGVHD status

| Days Post HSCT | Sacin Z-score Change* | | | | p value |
|---|---|---|---|---|---|
| | cGVHD Group | | No-cGVHD Group | | |
| | Mean | S.E. | Mean | S.E. | |
| 90 | 0.78 | 0.25 | 0.32 | 0.19 | 0.158 |
| 180 | 1.26 | 0.38 | 0.15 | 0.28 | 0.028 |
| 360 | 1.73 | 0.33 | 0.17 | 0.23 | 0.001 |

Footnote: *values represent the mean absolute change in $S_{acin}$ Z-score from baseline. Welch comparison; S.E., standard error.

**Table E11.** Absolute change in LCI between groups stratified by cGVHD status

| Days Post HSCT | LCI Z-score Change* | | | | p value |
|---|---|---|---|---|---|
| | cGVHD Group | | No-cGVHD Group | | |
| | Mean | S.E. | Mean | S.E. | |
| 90 | 0.44 | 0.29 | 0.07 | 0.32 | 0.390 |
| 180 | 1.00 | 0.38 | -0.22 | 0.28 | 0.016 |
| 360 | 1.36 | 0.54 | -0.01 | 0.47 | 0.065 |

Footnote: *values represent the mean absolute change in LCI Z-score from baseline. Welch comparison; S.E., standard error.

**Table E12.** Absolute change in $FEV_1$ between groups stratified by cGVHD status

| Days Post HSCT | $FEV_1$ Z-score Change* | | | | p value |
|---|---|---|---|---|---|
| | cGVHD Group | | No-cGVHD Group | | |
| | Mean | S.E. | Mean | S.E. | |
| 90 | -0.03 | 0.11 | -0.05 | 0.15 | 0.920 |
| 180 | -0.21 | 0.18 | -0.41 | 0.31 | 0.575 |
| 360 | -0.38 | 0.20 | -0.25 | 0.22 | 0.667 |

Footnote: *values represent the mean absolute change in $FEV_1$ Z-score from baseline. Welch comparison; S.E., standard error.

##### 3.4 Comparing Change in Lung Function Indices between Groups Stratified by cGVHD severity grade

**Table E13.** Comparison of absolute change in $S_{acin}$ Z-scores between groups stratified by NIH cGVHD grade

| Multiple Comparisons | | Mean Difference (X-Y)* | S.E. | p value |
|---|---|---|---|---|
| **NIH Grade (X)** | **NIH Grade (Y)** | | | |
| **1** | **0** | 0.84 | 0.71 | 0.64 |
| | **2** | -0.51 | 0.70 | 0.89 |
| | **3** | -3.05 | 0.97 | 0.01 |
| **2** | **0** | 1.35 | 0.69 | 0.22 |
| | **1** | 0.51 | 0.70 | 0.89 |
| | **3** | -2.53 | 0.95 | 0.048 |
| **3** | **0** | 3.89 | 0.96 | 0.001 |
| | **1** | 3.05 | 0.97 | 0.01 |
| | **2** | 2.53 | 0.95 | 0.048 |

Footnote: *Mean difference represents the mean difference in $S_{acin}$ Z-score change between NIH Grade in column (X) and NIH Grade in column (Y). Tukey's HSD post-hoc test; S.E., standard error.

Of the seven patients in the cGVHD grade 3 group, two had NIH defined BOS (stages 2 and 3) and two of the remaining five went on to develop BOS, soon after their 2nd HSCT. In these latter two patients, we only included data preceding their 2nd HSCT in the analysis. However, even within this follow-up period, both these subjects had detectable deteriorations in $S_{acin}$ of up to an absolute Z-score change of 3.7 SD from baseline values, in the absence of any deterioration in $FEV_1$. There were also two patients with BOS stage 1 in the cGVHD grade 2 group, but none with NIH defined lung involvement in the cGVHD grade 1 group.

##### 3.5 Ability to predict BOS-0p outcome

**Table E14**. Ability of early changes in $S_{acin}$ Z-score, from baseline, at 180 days to predict subsequent development of BOS-0p at 1, 2 and 3 years post-HSCT.

| Delta Sacin Z-score* | *1yr Post-HSCT* | | | | *2yrs Post-HSCT* | | | | *3yrs Post-HSCT* | | | |
|---|---|---|---|---|---|---|---|---|---|---|---|---|
| | **B** | **S.E.** | **Exp(B)** | **p-value** | **B** | **S.E.** | **Exp(B)** | **p-value** | **B** | **S.E.** | **Exp(B)** | **p-value** |
| ≥0.5 | 0.94 | 0.87 | 2.57 | 0.28 | 0.94 | 0.79 | 2.57 | 0.23 | 0.54 | 0.74 | 1.71 | 0.47 |
| ≥1.0 | 1.48 | 0.89 | 4.40 | 0.10 | 1.48 | 0.82 | 4.40 | 0.069 | 1.08 | 0.76 | 2.93 | 0.159 |
| ≥1.5 | 1.69 | 0.93 | 5.42 | 0.069 | 1.13 | 0.87 | 3.10 | 0.19 | 0.88 | 0.85 | 2.41 | 0.30 |
| ≥2.0 | 0.69 | 1.10 | 2.00 | 0.53 | 0.34 | 1.08 | 1.40 | 0.76 | 0.15 | 1.08 | 1.17 | 0.89 |

Footnote: *Delta $S_{acin}$ Z-score thresholds represent absolute Z-score change from baseline. Logistic regression; S.E., standard error.

**Table E15.** Change in MBW parameters Z-scores at 90 days, 180 days and 360 days vs. BOS-0p status at 1, 2 and 3 years post-HSCT

| | **BOS-0p Diagnosis** | | | | | | | | |
|---|---|---|---|---|---|---|---|---|---|
| | **1yr** | | | **2yrs** | | | **3yrs** | | |
| **Parameter Change** | **AUC** | **S.E.** | **p-value** | **AUC** | **S.E.** | **p-value** | **AUC** | **S.E.** | **p-value** |
| **90 Days Post-HSCT** | | | | | | | | | |
| Sacin Z-score | 0.467 | 0.154 | 0.817 | 0.479 | 0.113 | 0.850 | 0.462 | 0.108 | 0.722 |
| Scond Z-score | 0.508 | 0.135 | 0.954 | 0.592 | 0.103 | 0.406 | 0.595 | 0.100 | 0.374 |
| LCI Z-score | 0.567 | 0.148 | 0.644 | 0.542 | 0.112 | 0.705 | 0.549 | 0.107 | 0.644 |
| FEV1 Z-score | 0.550 | 0.151 | 0.729 | 0.467 | 0.116 | 0.762 | 0.428 | 0.113 | 0.500 |
| FEF25-75 Z-score | 0.400 | 0.148 | 0.488 | 0.400 | 0.122 | 0.364 | 0.417 | 0.115 | 0.434 |
| **180 Days Post-HSCT** | | | | | | | | | |
| Sacin Z-score | 0.536 | 0.143 | 0.785 | 0.545 | 0.126 | 0.702 | 0.522 | 0.117 | 0.846 |
| Scond Z-score | 0.286 | 0.112 | 0.101 | 0.338 | 0.124 | 0.171 | 0.412 | 0.114 | 0.438 |
| LCI Z-score | 0.536 | 0.141 | 0.785 | 0.552 | 0.106 | 0.661 | 0.511 | 0.117 | 0.923 |
| FEV1 Z-score | **0.143** | **0.087** | **0.006** | **0.227** | **0.106** | **0.021** | **0.220** | **0.098** | **0.013** |
| FEF25-75 Z-score | **0.134** | **0.103** | **0.005** | **0.240** | **0.113** | **0.029** | **0.236** | **0.100** | **0.020** |
| **360 Days Post-HSCT** | | | | | | | | | |
| Sacin Z-score | 0.706 | 0.112 | 0.101 | 0.685 | 0.105 | 0.111 | 0.072 | 0.103 | 0.089 |
| Scond Z-score | 0.627 | 0.122 | 0.313 | 0.589 | 0.114 | 0.440 | 0.599 | 0.111 | 0.382 |
| LCI Z-score | 0.746 | 0.109 | 0.051 | 0.708 | 0.105 | 0.072 | 0.714 | 0.101 | 0.058 |
| FEV1 Z-score | **0.056** | **0.051** | **<0.001** | **0.137** | **0.075** | **0.002** | **0.132** | **0.071** | **0.001** |
| FEF25-75 Z-score | **0.127** | **0.076** | **0.003** | **0.137** | **0.072** | **0.002** | **0.159** | **0.076** | **0.003** |

Footnote: S.E., standard error.

##### 3.6 Ability to predict cGVHD outcome

**Table E16.** Change in MBW parameters Z-scores at 90 days, 180 days and 360 days vs. cGVHD status at 1, 2 and 3 years post-HSCT

| | cGVHD Diagnosis | | | | | | | | |
|---|---|---|---|---|---|---|---|---|---|
| | 1yr | | | 2yrs | | | 3yrs | | |
| **Parameter Change** | **AUC** | **S.E.** | **p-value** | **AUC** | **S.E.** | **p-value** | **AUC** | **S.E.** | **p-value** |
| **90 Days Post-HSCT** | | | | | | | | | |
| Sacin Z-score | 0.615 | 0.109 | 0.312 | 0.632 | 0.102 | 0.217 | 0.617 | 0.102 | 0.271 |
| Scond Z-score | 0.594 | 0.11 | 0.41 | 0.617 | 0.101 | 0.277 | 0.621 | 0.1 | 0.256 |
| LCI Z-score | 0.545 | 0.115 | 0.689 | 0.542 | 0.11 | 0.699 | 0.542 | 0.11 | 0.696 |
| FEV1 Z-score | 0.476 | 0.116 | 0.832 | 0.506 | 0.109 | 0.956 | 0.489 | 0.108 | 0.915 |
| FEF25-75 Z-score | 0.358 | 0.109 | 0.213 | 0.427 | 0.105 | 0.496 | 0.436 | 0.104 | 0.546 |
| **180 Days Post-HSCT** | | | | | | | | | |
| Sacin Z-score | 0.684 | 0.089 | 0.063 | 0.676 | 0.088 | 0.065 | 0.677 | 0.088 | 0.062 |
| Scond Z-score | 0.575 | 0.092 | 0.444 | 0.618 | 0.085 | 0.218 | 0.622 | 0.083 | 0.198 |
| LCI Z-score | **0.738** | **0.079** | **0.016** | **0.72** | **0.077** | **0.021** | **0.736** | **0.074** | **0.013** |
| FEV1 Z-score | 0.43 | 0.097 | 0.479 | 0.438 | 0.096 | 0.518 | 0.442 | 0.096 | 0.539 |
| FEF25-75 Z-score | 0.499 | 0.097 | 0.988 | 0.513 | 0.094 | 0.893 | 0.525 | 0.092 | 0.79 |
| **360 Days Post-HSCT** | | | | | | | | | |
| Sacin Z-score | **0.8** | **0.096** | **0.016** | **0.794** | **0.088** | **0.012** | | | |
| Scond Z-score | 0.5 | 0.125 | 1 | 0.547 | 0.115 | 0.688 | | | |
| LCI Z-score | 0.731 | 0.108 | 0.063 | 0.718 | 0.1 | 0.063 | | NA | |
| FEV1 Z-score | 0.385 | 0.125 | 0.352 | 0.418 | 0.116 | 0.482 | | | |
| FEF25-75 Z-score | 0.5 | 0.124 | 1 | 0.547 | 0.112 | 0.688 | | | |

Within the cohort contributing for the 90 day analysis, 24/35 developed cGVHD during the follow up period and the mean±SD cGVHD onset was 315±258 days (range 100-1105). No subjects developed cGVHD before the 90 day threshold, which is to be expected as cGVHD only develops after this time point by definition (termed as acute GVHD if develops before).

Within this cohort contributing for the 180 day analysis, 35/48 developed cGVHD during the follow up period and the mean±SD cGVHD onset was 290±243 days (range 100-1105). 17/35 (49%) subjects developed cGHVD before the 180 day threshold.

Within this cohort contributing for the 360 day analysis, 17/27 developed cGVHD during the follow up period and the mean±SD cGVHD onset was 228±140 days (range 100-635). 16/17 (49%) subjects developed cGHVD before the 360 day threshold.
